# Functional and Computational Interrogation of the Juvenile Idiopathic Arthritis Risk Loci Identifies Candidate Causal SNPs and Target Genes in CD4+ T cells

**DOI:** 10.64898/2025.12.15.25342296

**Authors:** Kaiyu Jiang, Emma K. Haley, Gilad Barshad, Adam He, Anita Rogic, Edward J. Rice, Marc Sudman, Susan D Thompson, Charles G. Danko, James N. Jarvis

## Abstract

GWAS have identified multiple genetic regions that confer risk for juvenile idiopathic arthritis (JIA). However, identifying the single nucleotide polymorphisms (SNPs) that drive disease risk has been impeded by the fact that the SNPs used to identify risk loci are in linkage disequilibrium (LD) with hundreds of other SNPs. Since the causal SNPs remain unknown, it is difficult to identify target genes and thus use genetic information to elucidate disease biology and inform patient care.

We next used existing genotyping data from 3,939 children with JIA and 14,412 healthy controls to identify SNPs on JIA risk haplotypes that: present within open chromatin in multiple immune cell types and more common in children with JIA than the controls (p<0.05) in the genotyping data sets. We identified SNPs within cis-regulatory regions (CREs) using precision run-on sequencing data, and identified likely target genes using MicroC in both resting and activated CD4+ T cells. We identified 138 SNPs within the PROseq-identified CREs, and n=41 genes with which these CREs physically interacted. Data from GTEx corroborated these analyses by showing allelic effects for SNPs within the CREs in the *ERAP2* and *IRF1* risk loci. We further corroborated *IRF1* allelic effects using a luciferase reporter assay. Our findings significantly reduce the genomic search space for risk-driving variants and target genes and support the roles of *IRF1, ERAP2* and *LNPEP* in driving risk for JIA.

## Introduction

Juvenile idiopathic arthritis (JIA) is a term used to describe a group of childhood illnesses characterized by chronic inflammation and hypertrophy of synovial membranes. JIA is one of the most common chronic disease conditions in children^1,2^, and, although treatment outcomes have improved over the past 20 years, true remission is rare^3^; more than 50% of adults who had JIA continue to have active disease^4^.

JIA has long been recognized as a complex genetic trait, in which multiple genetic loci contribute to disease risk^5^. Although the contribution of any single genetic locus is small, genetic influences exert a significant effect, as shown by family studies using the using the Utah Population Database^6^. More than 30 different risk loci have been suggested to contribute to JIA, ^7–9^ including those derived from the Hinks Immunochip study^8^ and the McIntosh and Lopez-Isacs meta-analyses^9,10^. We have shown that the JIA risk loci are highly enriched, compared to genome background, for H3K27me1 and H3K27ac ChIPseq peaks, epigenetic features typically associated with enhancer function^11–13^. In this regard, JIA resembles multiple other complex traits, including autoimmune diseases^14,15^. Furthermore, using *in* vitro approaches, we have demonstrated that variants on the JIA risk haplotypes alter the efficiency of enhancers within the *IL2RA* and *IL6R* risk loci ^16^.

The presence of suitable chromatin features (e.g., open chromatin and abundant transcription factor binding between H3K27ac ChIPseq peaks) is not, in itself, *prima facie* evidence that a region has enhancer activity. In order to better define the regions likely most likely to harbor the disease-driving SNPs on JIA risk haplotypes, additional information may be necessary to identify the specific non-coding functional elements likely to harbor (and be impacted by) those variants. One of the key features of active enhancers is the presence of bidirectional RNA synthesis, which can be detected by techniques like global run-on sequencing (GROseq) and precision run-on sequencing (PROseq) ^17^. In this paper, we used multiple genomic data sets to refine the list of potential candidate causal SNPs within the JIA-associated LD blocks and the genes likely to be influenced by those SNPs. Because of the emerging importance of interferon-regulated pathways in our understanding of JIA^18^, We also performed additional functional studies to elucidate genetic mechanisms within the *IRF1* risk locus.

## Methods

### Genotyping data sets

We queried raw data from genome-wide association studies and genetic fine mapping studies as previously reported in^8,9^. From McIntosh et al. three cohorts comprising 2,751 patients with oligoarticular or RF-negative polyarticular JIA were genotyped using the Affymetrix Genome-Wide SNP Array 6.0 or the Illumina HumanCoreExome-12+ Array. Overall, 15,886 local and out-of-study controls, typed on these platforms or the Illumina HumanOmni2.5, were used for association analyses.

From Hinks et al. Immunochip array was used to analyze 2,816 individuals with juvenile idiopathic arthritis (JIA), comprising the most common subtypes (oligoarticular and rheumatoid factor-negative polyarticular JIA), and 13,056 controls. Combined, these studies include genotyping data from 3,939 children with oligoarticular/polyarticular (RF-negative) JIA and 14,412 healthy controls. Using r^2^ >0.50, we identified all SNPs on those haplotypes listed in **Table 1** that were in linkage disequilibrium (LD) with the tag SNPs. We further filtered these SNPs using the QC analysis methods described in^8,9^ (e.g., presence of Hardy Weinberg equilibrium, minor allele frequency, missingness), and identified those that were significantly more common in patients than in controls (p <0.05). Finally, from this list, we identified SNPs that were situated within regions of open chromatin and within H3K4me1/H3K27ac peaks in CD4+ T cells^12^. These filtering processes provided us with a list of 846 candidate SNPs.

**Table 1:** Positional information of the 23 JIA-risk single nucleotide polymorphisms and the associated haplotypes.

| <u>Locus Name</u> | <u>Tag SNP</u> | <u>Hg38 Haplotype</u> |
| --- | --- | --- |
| PTPN22 | rs6679677 | chr1:113,761,186-113,834,946 |
| STAT4 | rs10174238 | chr2:191,079,016-191,108,308 |
| PTPN2 | rs2847293 | chr18:12,774,327-12,809,341 |
| ANKRD55 | rs71624119 | chr5:56,141,024-56,146,422 |
| IL2-IL21 | rs1479924 | chr4:122,151,854-122,619,603 |
| TYK2 | rs34536443 | chr19:10,317,045-10,381,598 |
| IL2RA | rs7909519 | chr10:6,028,313-6,055,320 |
| SH2B3-ATXN2 | rs3184504 | chr12:111,395,984-111,645,358 |
| ERAP2-LNPEP | rs27290 | chr5:96,884,383-97,038,046 |
| UBE2L3 | rs2266959 | chr22:21,556,931-21,628,971 |
| C5orf56/IRF1 | rs4705862 | chr5:132,477,527-132,496,822 |
| RUNX1 | rs9979383 | chr21:35,323,611-35,365,944 |
| IL2RB | rs2284033 | chr22:37,135,077-37,141,474 |
| ATP8B2/IL6R | rs11265608 | chr1:154,319,242-154,406,893 |
| ZFP36L1 | rs12434551 | chr14:68,784,174-68-794,755 |
| JAK1 | rs10889504 | chr1:64,924,820-64,975,081 |
| PRR9/LOR | rs873234 | chr1:153,249,128-153,270,022 |
| PTH1R | rs1138518 | chr3:46,889,988-46,932,682 |
| ILDR1/CD86 | s111700762 | chr3:122,023,892-122,102,059 |
| AHI1/LINC00271 | rs9321502 | chr6:135,303,673-35,371,709 |
| HBP1 | rs111865019 | chr7:107,156,092-107,390,877 |
| WDFY4 | rs1904603 | chr10:48,776,493-48,805,795 |
| RNF215 | rs57533109 | chr22:30,289,571-30,403,731 |

### Defining LD blocks/JIA risk haplotypes

We queried 23 single nucleotide polymorphisms (SNPs) in non-HLA loci with established and replicated associations with JIA in the Hinks genetic fine mapping (Immunochip) study and the McIntosh meta-analysis of previous genome-wide scans ^8,9^. We then used the SNiPA online single nucleotide polymorphism annotator^19^ (https://snipa.helmholtz-muenchen.de/snipa3/) to define linkage disequilibrium (LD) blocks for each of the 23 SNPs. We used the following settings: GRCh37, 1000 Genomes Phase 3 v5, querying European populations, and setting r^2^ at 0.8. The smallest genomic position was used as the start of the LD block while the largest position was used as the end of the block. Although our initial set of n=846 SNPs was generated using r^2^ = 0.5, we focused this study on the regions identified using r^2^ = 0.80. We subsequently converted GRCh37/hg19 coordinates to GRCh38/hg38 using the liftover tool to map relevant chromatin data. These regions are shown in **Table 1**.

### Isolation of CD4+ T cells from peripheral blood

Peripheral blood samples (60–80 mL) were obtained from healthy adult donors under protocols approved by the Cornell University Institutional Review Board. Informed consent was obtained from all participants. Samples were collected from three independent donors. Blood was drawn into EDTA-coated collection tubes and stored overnight at 4°C to simulate potential shipping delays. Following dilution in an equal volume of phosphate-buffered saline (PBS), peripheral blood mononuclear cells (PBMCs) were isolated by centrifugation at 750 × g for 30 minutes at 20°C over a 15 mL layer of Ficoll-Paque. Isolated PBMCs were washed three times in ice-cold PBS. CD4+ T-cells were purified using human-specific CD4 microbeads (Miltenyi Biotech, 130-045-101). Up to 10⁸ PBMCs were resuspended in binding buffer (PBS supplemented with 0.5% BSA and 2 mM EDTA) and incubated with 20 μL of CD4 microbeads per 10⁷ cells for 15 minutes at 4°C in the dark. After washing in PBS/BSA buffer, cells were resuspended in 500 μL binding buffer and applied to a MACS LS column (Miltenyi Biotech, 130-042-401) placed in a neodymium magnetic separator. Columns were washed three times with 2 mL PBS/BSA before removing from the magnet and eluting the bound CD4+ cells. Cell counts were obtained using a hemocytometer.

### CD4+ T cell stimulation

Purified CD4+ T-cells were allowed to rest in RPMI-1640 medium supplemented with 10% fetal bovine serum for 2–4 hours prior to stimulation. Cells were treated for 30 minutes with either vehicle control (2.5 μL ethanol and 1.66 μL DMSO per 10 mL medium) or with 25 ng/mL phorbol 12-myristate 13-acetate (PMA) and 1 μM ionomycin as described in ^20^. Cells were lysed in 1 mL of ice-cold lysis buffer (10 mM Tris-Cl pH 8.0, 300 mM sucrose, 10 mM NaCl, 2 mM MgAc₂, 3 mM CaCl₂, and 0.1% NP-40) to release intact nuclei.

Nuclei were washed in 10 mL of wash buffer (10 mM Tris-Cl pH 8.0, 300 mM sucrose, 10 mM NaCl, 2 mM MgAc₂) to remove unincorporated nucleotides, then pelleted and resuspended in 1 mL and finally 50 μL of storage buffer (50 mM Tris-Cl pH 8.3, 40% glycerol, 5 mM MgCl₂, and 0.1 mM EDTA). Nuclei were snap frozen in liquid nitrogen and stored at –80°C for up to 6 months prior to PRO-seq library preparation.

For longer term stimulation, we cultured CD4+ T cells in expansion media (ImmunoCult-XF T Cell Exp Medium, STEMCELL Technologies Inc.; 10ng/ml IL2) at 1 x 10^6^/ml, at 37°C, 5% CO2.

We activate cells for 3 days using human CD3/CD28/CD2 T cell activator (STEMCEL Technologies, Inc.), followed by culture in expansion media (adjusted to 1.25×10^6^/ml) for 3 days without antibiotics. Cells were harvested by centrifuge for 10 minutes at 400x*g* and frozen in liquid nitrogen until used for PROseq and/or MicroC studies.

### Assessing functional regions: dREG PROseq in resting and activated CD4+ T cells

We created PRO-seq libraries following the method outlined by Judd et al^21^. The process was started by performing a run-on reaction with four biotin nucleotides. The nascent RNA was then purified, ligated to the 3’ adapter, and bound to streptavidin beads and washed. The 5’ cap was removed with RppH, then the 5’ end is phosphorylated. We then ligated a 5’ adapter to the RNA transcript, and purified the RNA from the streptavidin beads. Following these steps, a reverse transcription reaction was used to generate cDNA, which was amplified by PCR. Final PRO-seq libraries sequenced using either an Illumina Hi-Seq 2000, NextSeq 500, or NovaSeq X series instrument. PRO-seq data was mapped to the reference genome using established pipelines^22^. BigWig files were uploaded to the web-based dREG Gateway for analysis^4^.

### MicroC analyses

We used MicroC ^23,24^ to identify genes whose promoters interact with regulatory regions harboring the candidate causal SNPs within the JIA risk regions. We prepared CD4+ T cells as described above and crosslinked cells with1% formaldehyde, quenching reactions using 0.25 M Glycine for 5 min. After spin-down for 5 minutes at 300Xg at 4 °C, cells were washed at a density of 1 ml per million cells in ice cold PBS. Cells were crosslinked a second time, with 1 ml per 4 million cells of 3 mM disuccinimidyl glutarate (DSG) (ThermoFisher Scientific, 20593) for 40 min at room temperature and quenched by 0.4 M Glycine for 5 min. Following washes with cold PBS, cells were flash-frozen cells and stored at - 80°C.

For MNase digestion, cells were thawed on ice for 5 min, incubated with 1ml MB#1 buffer (10 mM Tris-HCl, pH 7.5, 50 mM NaCl, 5 mM MgCl2, 1 mM CaCl2, 0.2% NP-40, 1x Roche cOmplete EDTA-free (Roche diagnostics, 04693132001)) and washed with MB#1 buffer. We determined MNase concentrations using MNase titration experiments exploring 2.5-20U of MNase per million cells. Chromatin was digested with MNase for 10 min at 37 °C and digestion stopped by adding 500 mM EGTA and incubating at 65 °C for 10 min. Following dephosphorylation with rSAP and end polishing using T4 PNK, DNA polymerase Klenow fragment (NEB #M0210) and biotinylated dATP and dCTP, we performed ligation in a final volume of 2.5 ml for 3h at room temperature using T4 DNA ligase. Dangling ends were removed by a 5 min incubation with Exonuclease III (NEB #0206) at 37 °C and biotin enrichment was done using 20 ul DynabeadsTM MyOneTM Streptavidin C1 beads. We prepared libraries with the NEBNext Ultra II Library Preparation Kit (NEB #E7103), and sequenced samples on a combination of Illumina’s NovaSeq 6000 and HiSeq 2500 at Novogene to a depth of 1 billion mapped reads combined across all samples.

### Contact enrichment analysis

We identified interactions between dREG regions containing JIA risk haplotypes and the candidate target genes using the Micro-C contact caller (MCC)^25,26^. Micro-C contacts that were mapped in one end to a 5kb window around the centers of dREG sites that overlapped with JIA risk associated SNPs and on the other end to a 5kb window around the transcription start site (TSS) of genes within 1Mbp from the center were be captured. We used a LOWESS regression-based contact caller^27^ to estimate enrichment of contacts relative to the expected given the contacts-by-distance decay in regions of up to 1Mbp from the center of the dREG peak – at the TSS orientation. We used Fisher’s exact p-values corrected by Benjamini-Hochberg false discovery rate (BH FDR) of 0.1 as a cutoff for a significant interaction between the JIA risk-associated SNP overlapping dREG site and a gene TSS.

### Identification of dREG PRO-seq peaks within JIA LD risk blocks

We next used the dREG web gateway to query PROseq data to identify functional cis-regulatory elements (CREs) in each CD4+ T cell dataset. In the 23 risk haplotypes, we assessed the presence of CREs, which identify divergent transcription characteristic of active transcription initiation, within the 23 risk regions. We used the intersect command within the BEDTools software package to identify intersections between dREG PRO-seq peak data and these linkage disequilibrium LD regions. To determine significance, we created 23 random regions with length equal to the average length of the 23 LD blocks of interest (144,134 bp). Using bedtools intersect, we determined the number of random regions that overlapped with the dREG PRO-seq peak file. We repeated this process 1000 times to approximate a normal distribution. We then determined where the number of overlaps with the regions of interest fell within the normal curve in order to calculate the associated p value. We considered a Benjamini Hochberg false discovery rate corrected *p* value of *p*<0.1 as statistically significant.

### Luciferase enhancer reporter assay

As proof of concept that alleles within the JIA risk loci can alter enhancer function within primary human CD4+ T cells, we performed enhancer reporter assays. These assays are described in detail in^13^, and are described briefly here. A 706bp DNA sequence (chr5:132,496,614-132,497,319) was generated by PCR (Primers: <u>F:5’-TTTCCGCGAACACCTCTG-3’ and R:5’-ACTCCTCCCTCATTCCCTAAA-3</u>’) XhoI (5′-C^TCGAG-3′) and BglII (5′-A^GATCT-3′) restriction endonuclease recognition sites were incorporated in the oligonucleotides for directional cloning. The PCR amplified fragments were digested using XhoI and BglII (New England Biolabs, MA, USA) and ligated into a pGL4.23[*luc2*/minP] vector (Promega E8411, Madison, MI, USA), previously digested using the same endonucleases. The ligation mixtures were transformed into Invitrogen™ One Shot™ TOP10 Chemically Competent E. coli. Plasmid DNA was isolated by Miniprep kit (Qiagen) according to the manufacturer’s instructions.

Mutations were introduced with the QuikChange II Site-Directed Mutagenesis Kit (Agilent, Santa Clara, CA). The wild-type (wt) construct pGL4.23-IRF1 enhancer (chr5:132,496,614-132,497,319) was used as template to generate enhancer genetic variant. The primer oligonucleotide sequences were: Forward: <u>5’-CCGCTCAGCACTACCCGCGCCTC-3’ and Reverse: 5’-GAGGCGCGGGTAGTGCTGAGCGG-3’.</u> All recombinant plasmid sequences were confirmed by Sanger sequencing.

Cells were transiently transfected using electroporation following the protocol of the manufacturer (Nucleofector Device with Cell line kit V, Lonza, USA). Activated human CD 4^+^ T cells (5×10^6^) were transfected with 1.5 ug of the pGL4.23-constructs plus 1.5 ug of the control Renilla plasmid DNA (pRL-TK; Promega) to normalize for transfection efficiency. Twenty-four hours after transfection, the cells were harvested, lysed and analyzed for luciferase activity.

Luminescence was measured with the Dual-Luciferase^®^ Reporter Assay System (Promega) in a microplate luminometer (GloMax® Discover Microplate Reader, Promega). Results from each construct and from the empty vector were normalized by dividing the luciferase reporter activity by the renilla reporter activity. Relative luciferase activities were calculated by dividing each normalized construct by the normalized negative control (empty vector). Relative luciferase activities are representative of four independent transfection experiments. Student’s t test was used to determine statistical significance.

*Corroboration of SNPs and Gene Targets Using Genotype-Tissue Expression (GTEx) Project Data* - After identifying SNPs within PROseq-dREG peaks and their likely target genes, we next sought further corroboration that the identified SNPs influence the putative target genes. Using the GTEx eQTL calculator (https://www.gtexportal.org/home/testyourown), we queried expression data for the genes of interest for 3 cell types/tissues: whole blood, EBV-transformed lymphocytes, and spleen.

## Results

### JIA risk loci are enriched for PROseq peaks

On the 23 JIA risk haplotypes of interest, we used dREG to identify a total of 121 candidate CREs on 19 of the haplotypes in resting T cells (**table xxx**). These peaks are located within both intronic and intergenic regions. Note that a small portion of these peaks appear in exons, a finding consistent with the idea that some exons also have enhancer activity^28^. We performed enrichment analysis to determine whether the JIA risk haplotypes were more likely that random regions of the genome to fall in CD4+ T cell CREs.

We found strong enrichment (compared to genome background) for CREs in CD4+ T cells on the JIA risk haplotypes (p<0.0001). Our findings corroborate our previously published work, which showed enrichment for H3K4me1/H3K27ac ChIPseq peaks in these regions^11,12^. **(Supplemental Tables 1-3).**

### Differences in enrichment for PROseq peaks in the JIA risk loci between resting and activated T cells

On the 23 JIA risk haplotypes of interest, we identified the total number of CREs on the haplotypes in both resting and activated CD4+ T cells. **(Table 2).**

**Table 2:** Number of PROseq peaks in JIA haplotypes for CD4+ T cell groups.

| <u>Resting/Activated T cells</u> | <u># PROseq peaks</u> | <u>Shared PROseq peaks with CD3+CD28+ IL-2 activated</u> | <u>Shared PROseq peaks with PMA activated</u> |
| --- | --- | --- | --- |
| Resting CD4+ | 121 | 92 | 79 |
| CD3+CD28+ IL-2 activated | 147 | - | 77 |
| PMA activated | 121 | - | - |

IL-2 activated T-cells had a higher enrichment than the resting T cells, and the nonspecific activation by PMA/lo resulted in similar number of CREs as in resting T cells, but the CREs were in fewer JIA haplotypes (**Table 3**).

**Table 3:**
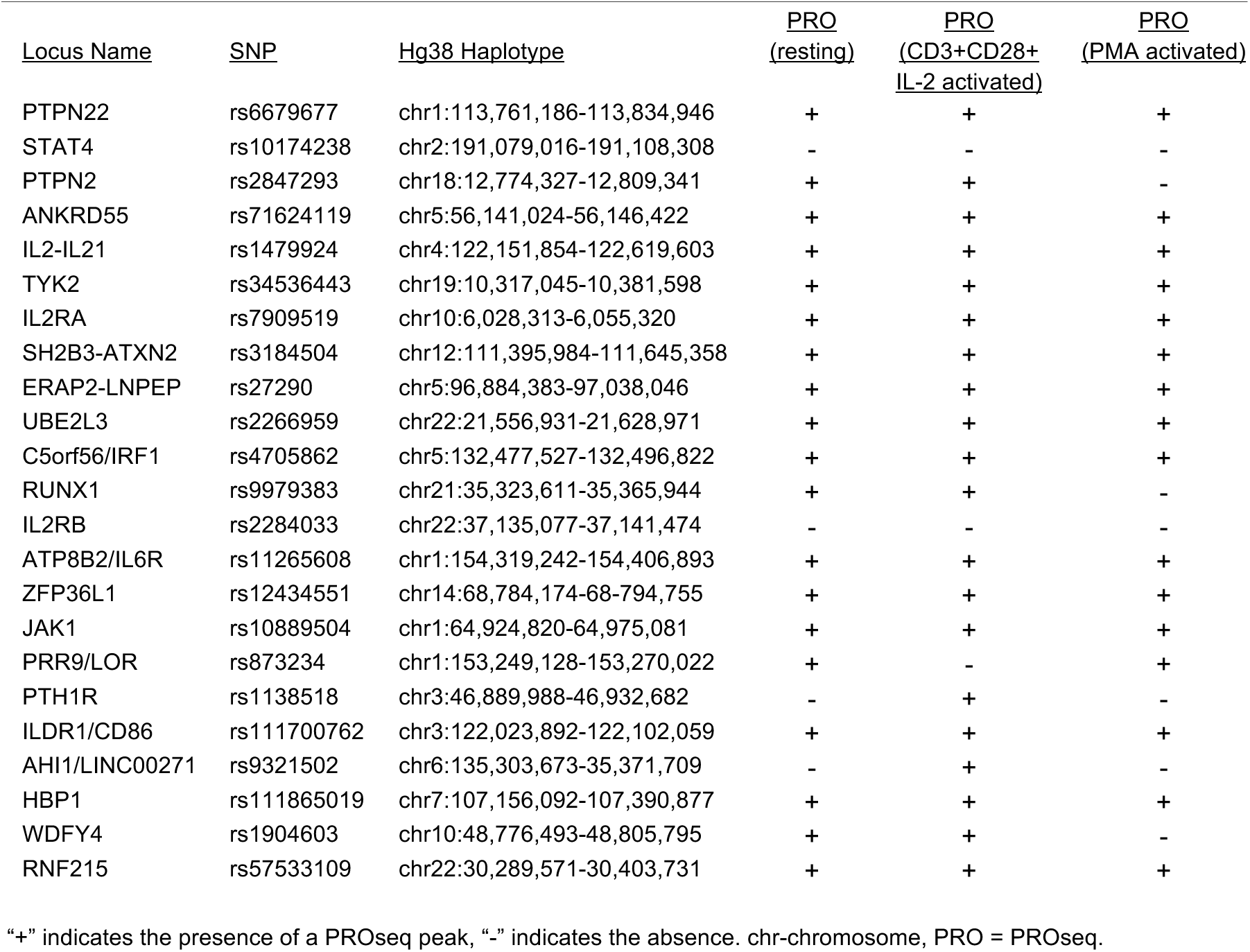
PROseq peaks that overlap the 23 JIA-risk SNPs and the associated haplotypes.

### Identification and characterization of SNPs located within PROseq-dREG CREs

Using our genotyping data, we identified n=138 SNPs situated within CREs in resting CD4+ T cells, 119 that occurred in CREs in PMA-stimulated CD4+ T cells (2 hr stimulation), and 150 SNPs that occurred in CREs in CD3/CD28/CD2-stimulated CD4+ T cells (5 days’ stimulation). We identified n=74 SNPs that appeared in CREs under all 3 conditions, suggesting that their biological effects are exerted on both resting and activated cells, while those unique to activation phases (20 in resting cells, 20 in PMA-activated cells, and 39 in CD3/CD28/IL2-activated cells) may exert their effects only under specific biological conditions. The list of SNPs within CREs for each condition is provided in **Supplemental Table 4.**

### Identification of target genes using Micro C

We next sought to identify the genes that are regulated by the CREs identified by PROseq-dReg analysis. Results are summarized in **Figure 1**, and **Figure 2** shows a representative heatmaps, from the *IRF1* and *ERAP2/LNPEP loci*. We identified n=41 genes that interacted with enhancers harboring the candidate causal SNPs in resting CD4+ T cells, n=39 in CD4+T cells activated with ionomycin and PMA (2 hr) and n=38 in CD4+ T cells activated with CD3/CD28/IL2 (5 days). There was considerable overlap in these gene sets, as shown in the **Figure 1**, while other gene-enhancer interactions were cell-state-specific. These findings, like the PROseq-dReg analyses, are consistent with findings by Gate et al ^29^ as well as our recently published MPRA study ^30^. demonstrating that the biological effects of risk-driving SNPs are likely to be exerted in specific immunologic contexts.

**Figure 1.**
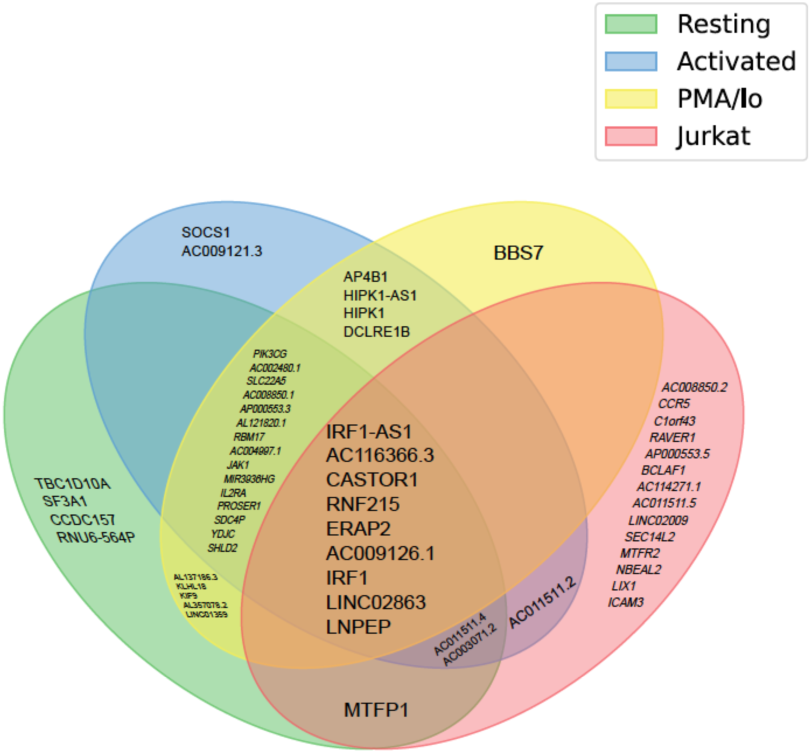
Venn diagram summarizing results of MicroC experiments elucidating genes whose promoters interact with PROseq-dREG regions harboring the candidate causal SNPs on JIA risk haplotypes. Data from unstimulated Jurkat cells are shown as a comparison. In primary human CD4+ T cells, there was considerable overlap between the candidate target genes identified is resting and activation CD4+ T cells. The largest differences were observed between (unstimulated) Jurkat cells and primary human CD4+ T cells, regardless of activation state.

**Figure 2.**
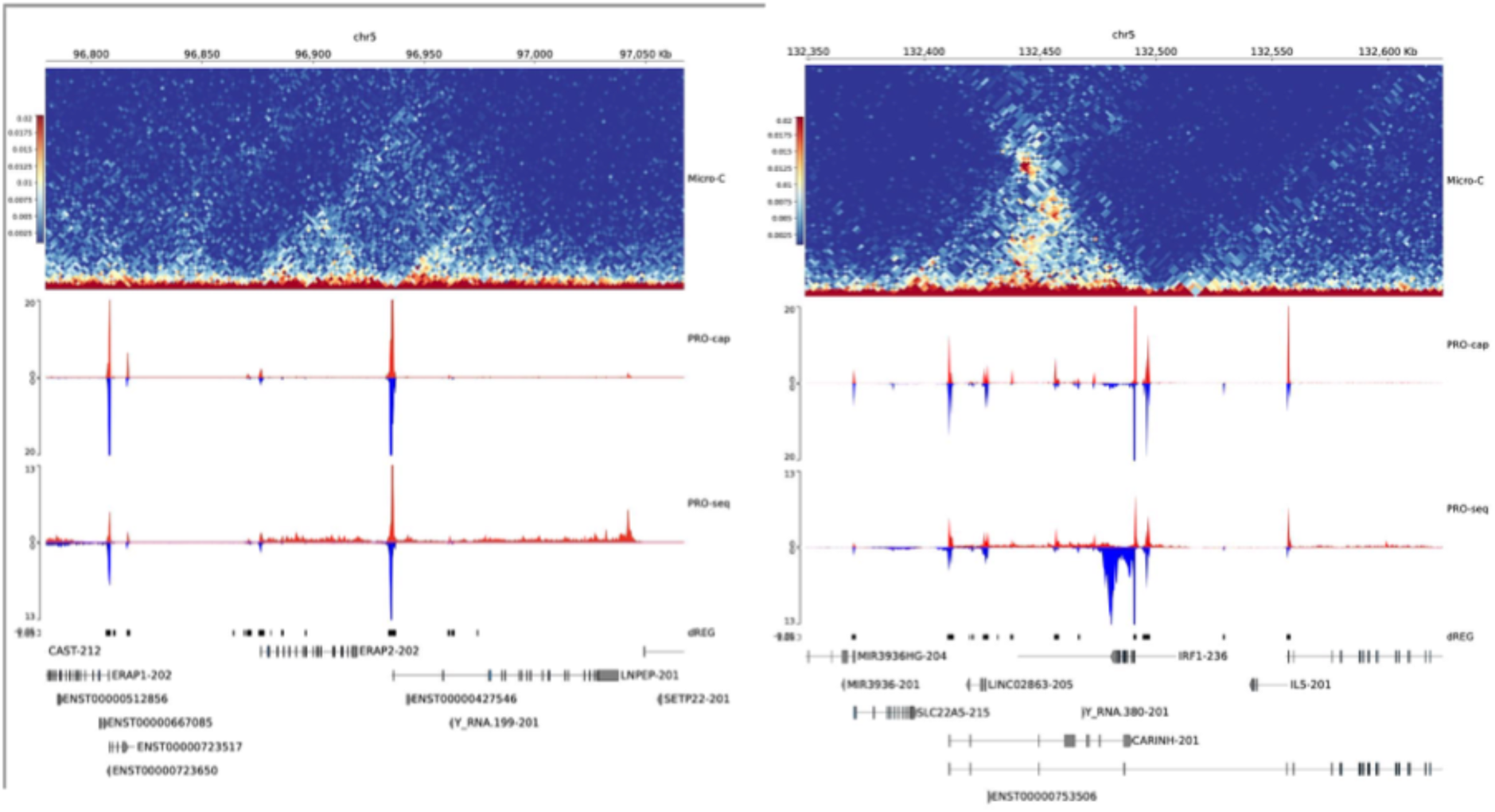
Heat maps derived from MicroC analysis of resting human CD4+ T cells. Shown are the regions encompassing ERAP2/LNPEP (left panel) and IRF1 (right panel). RNA PolII positioning, as detected by PROseq, is shown on the sense (red) and antisense (blue) strands. Positions of the candidate causal JIA SNPs is shown with the vertical black lines. Note that chromatin shows regions of contact forming micro-compartments (black and red triangles) that encompass the promoters of ERAP2 and LNPEP (left) and IRF1 (right). These findings are consistent with the presence of internal enhancers that regulate these genes.

### Corroboration Of Findings Using GTEx Data

We next sought to further evidence that the SNPs situated within the PROseq-dREG CREs influenced the candidate target genes identified by MicroC analyses. We focused on two regions/genes that were common to all 3 activation states in CD4+ T cells: *IRF1*and *ERAP2-LNPEP*. In the *IRF1* region, we identified two distinct CREs, one within the promoter region and another within an intergenic region upstream of the *IRF1* transcription start site. For each region, GTEx data demonstrated allele specific effects on expression of *IRF1* for SNPs within both of the CREs. **Figure 3** shows results from EBV-transformed lymphocytes (note that SNPs effects were sometimes associated with expression levels in other tissues as well). We also identified similar associations with gene expression for SNPs within the *LNPEP-ERAP2* locus. In this case, we also identified CREs within two different regions, one within an intronic region in the *ERAP2* gene and the other within an intronic region of the *LNPEP* gene. In this case, SNPs within each region demonstrated allele-specific effects on expression levels of both genes, as show in **Figures 4 and 5**. These analyses support the idea that individuals who carry specific alleles within the identified CREs display significant differences in expression of the candidate target genes.

**Figure 3.**
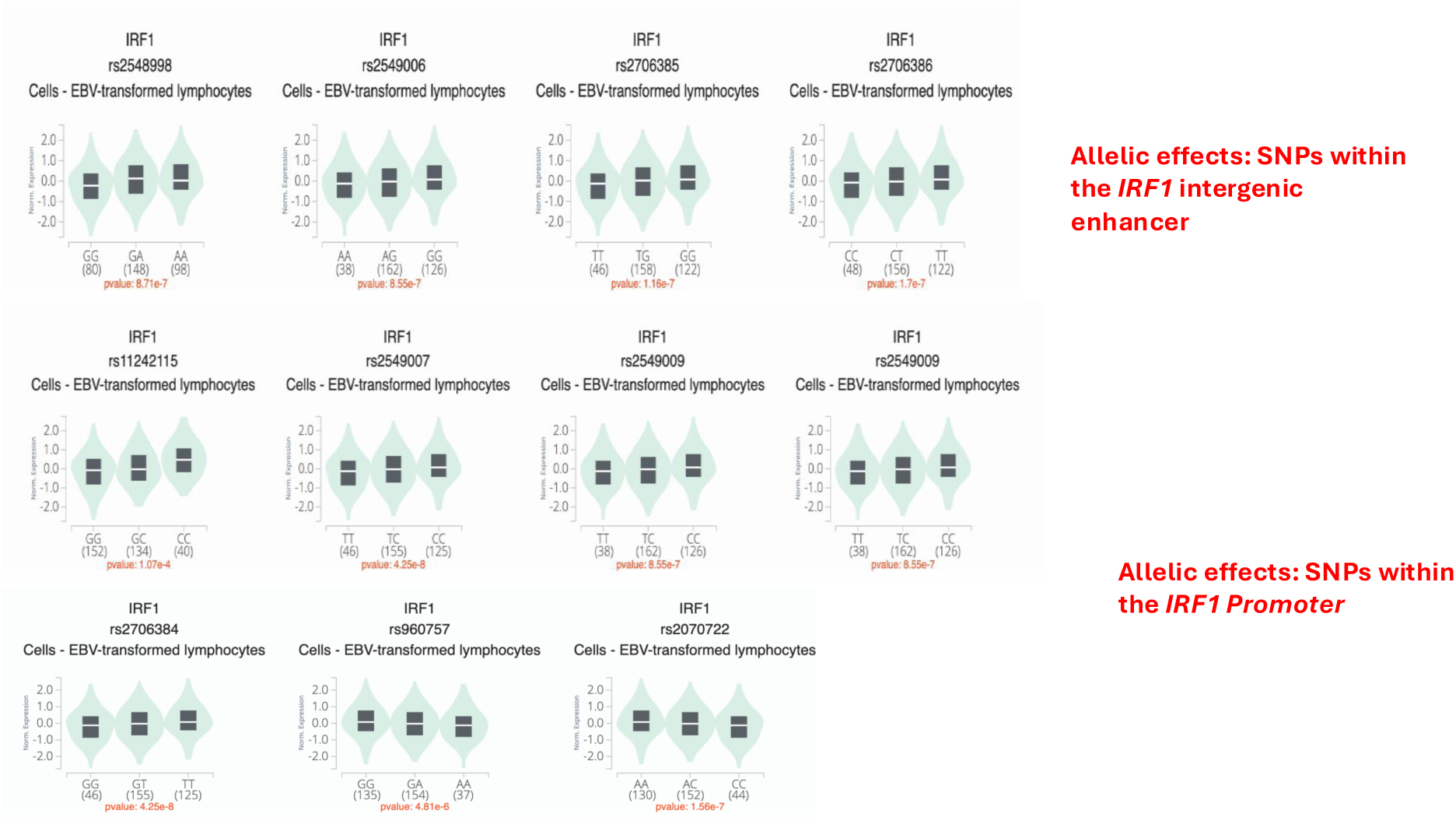
Violin plots showing allelic effects associated with IRF1 expression detected on eQTL analyses of GTEx data from EBV-transformed lymphocytes. The SNPs analyzed were situated within CREs situated within either an intergenic region upstream of the IRF1 transcription start site (top panel) or IRF1 promoter (bottom panel). P=values showing the likelihood of the association between the allele and expression of IRF1 are shown at the bottom of each violin plot.

**Figure 4.**
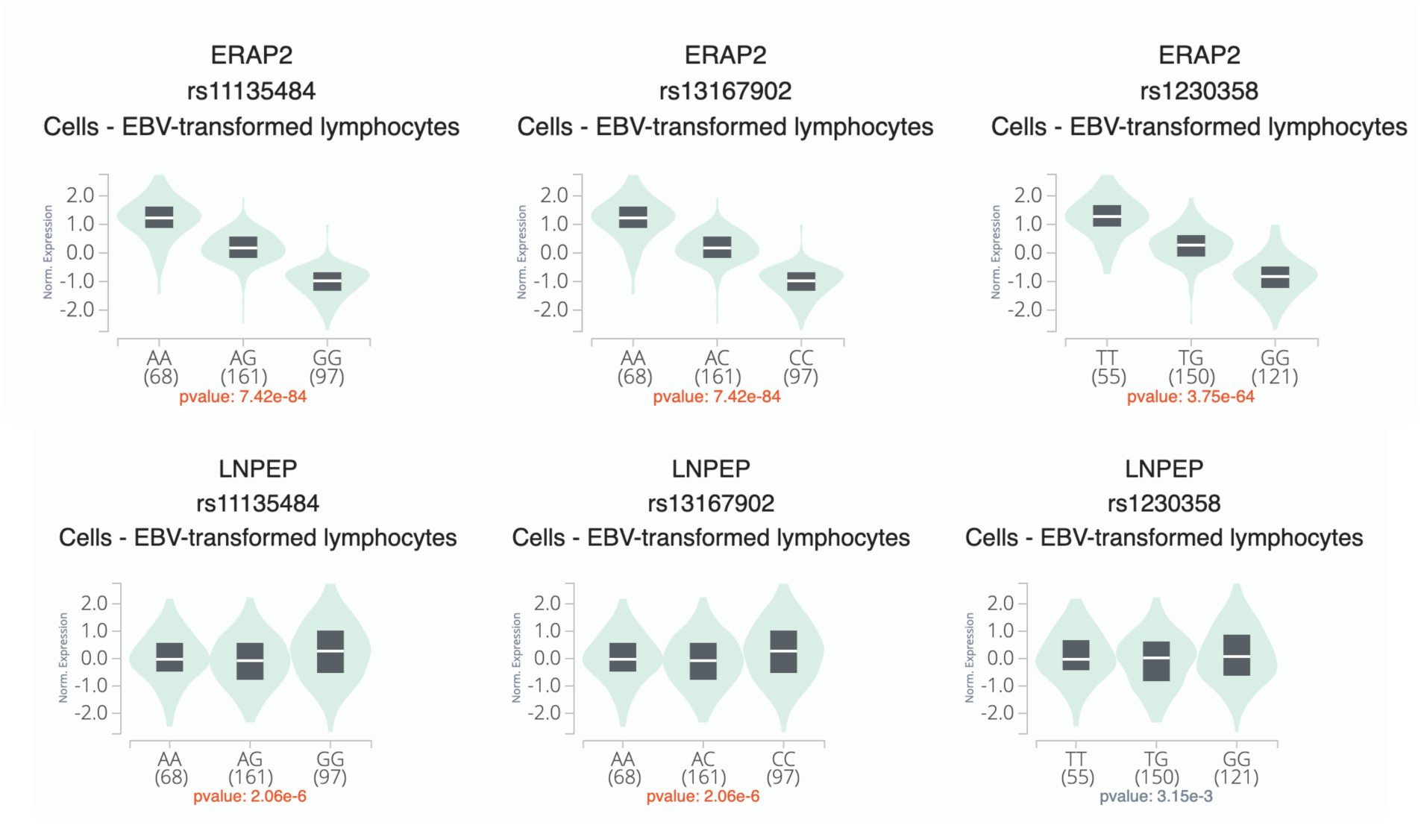
Violin plots summarizing allelic effects associated with ERAP2 (top panel) and LNPEP (bottom panel) expression detected on eQTL analyses of GTEx data. from EBV-transformed lymphocytes. The SNPs analyzed were situated within PROseq-dREG peaks situated within an intronic region within the ERAP2 gene. P=values showing the likelihood of the association between the allele and expression of ERAP2 or LNPEP are shown at the bottom of each violin plot. Note that the association is stronger for ERAP2 than LNPEP.

**Figure 5.**
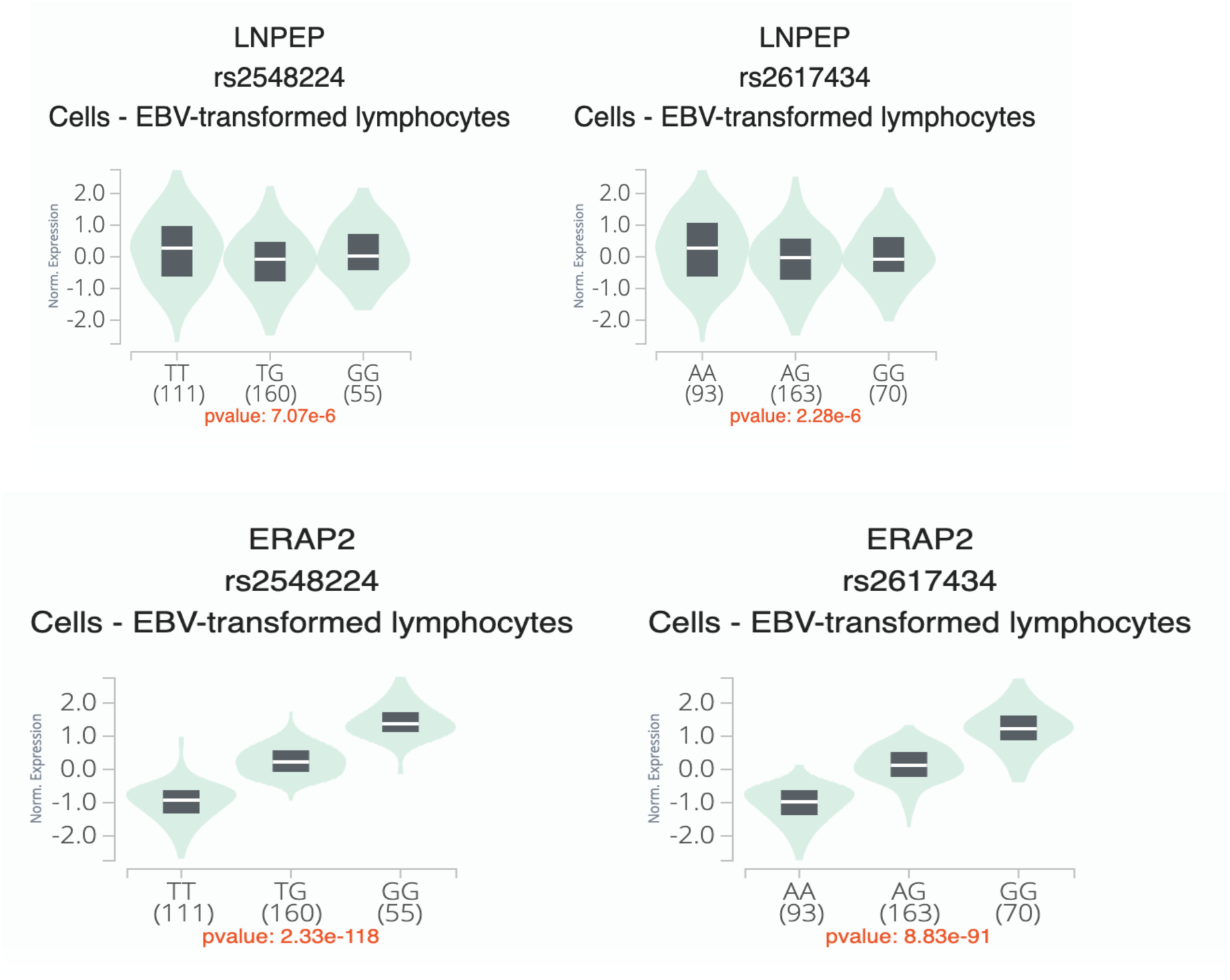
Violin plots summarizing allelic effects associated with LNPEP (top panel) and ERAP2 (bottom panel) expression detected on eQTL analyses of GTEx data from EBV-transformed lymphocytes. The SNPs analyzed were situated within PROseq-dREG peaks situated within an intronic region within the LNPEP gene. P=values showing the likelihood of the association between the allele and expression of ERAP2 or LNPEP are shown at the bottom of each violin plot. Note that, although the SNPs queried are situated within the LNPEP gene, the association is stronger for ERAP2 than LNPEP.

### Functional interrogation of the IRF1 locus

As noted in the methods section, we next sought further confirmation that the regions harboring PROseq peaks act as functional enhancers. We have previously shown that intronic regions in *IL2RA*, and an intergenic region in *ATP8B2/IL6R* have enhancer activity that can be altered by JIA-associated variants^13^, and that an intergenic enhancer that encompasses MPRA-screened alleles within the *LNPEP/ERAP2* locus has enhancer activity that regulates both *LNPEP* and *ERAP2*, but not the adjacent calpastatin (*CAST*) gene. Because of the prominence of genes regulated by interferons in data set (**Figure 1**), we undertook further functional analyses of the *IRF1* locus. This region includes a large enhancer complex marked by open chromatin and H3K27ac ChIPseq peaks in multiple immune cell types and ENCODE cell lines (**Figure 6**), spanning 3,967 bp from chr5:132,495,061-132,499,027. We identified a common variant rs2548998 G→A located in a candidate enhancer approximately 6kb upstream of the *IRF1* promoter as likely to influence transcription due to its position near an enhancer RNA TSS. Using a luciferase enhancer reporter assay, we demonstrated that the rs2548998 G allele showed 2x more activity than the A allele in activated primary human CD4+ T cells, as shown in **Figure 7**. The experimental finding is corroborated by GTEx data, which shows higher expression of *IRF1* in heterozygous individuals carrying the A allele, and the highest expression levels in AA homozygous individuals. Thus, while GTEx data are unable to disentangle LD to identify the SNPs/alleles that unambiguously drive expression, these experiments show that the alleles of rs2548998 have intrinsic ability to regulate gene expression independent of LD.

**Figure 6.**
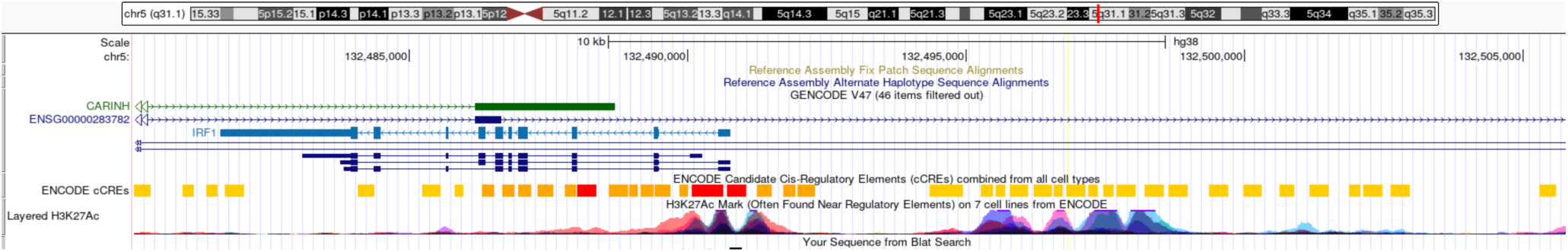
UCSC Genome Browser screen shot showing the JIA-associated IRF1 locus. Red-blue-maroon peaks are derived from ENCODE H3K27ac ChIPseq data. These peaks overlap PROseq-dREG peaks identified in CD4+ T cells (resting and activated).

**Figure 7.**
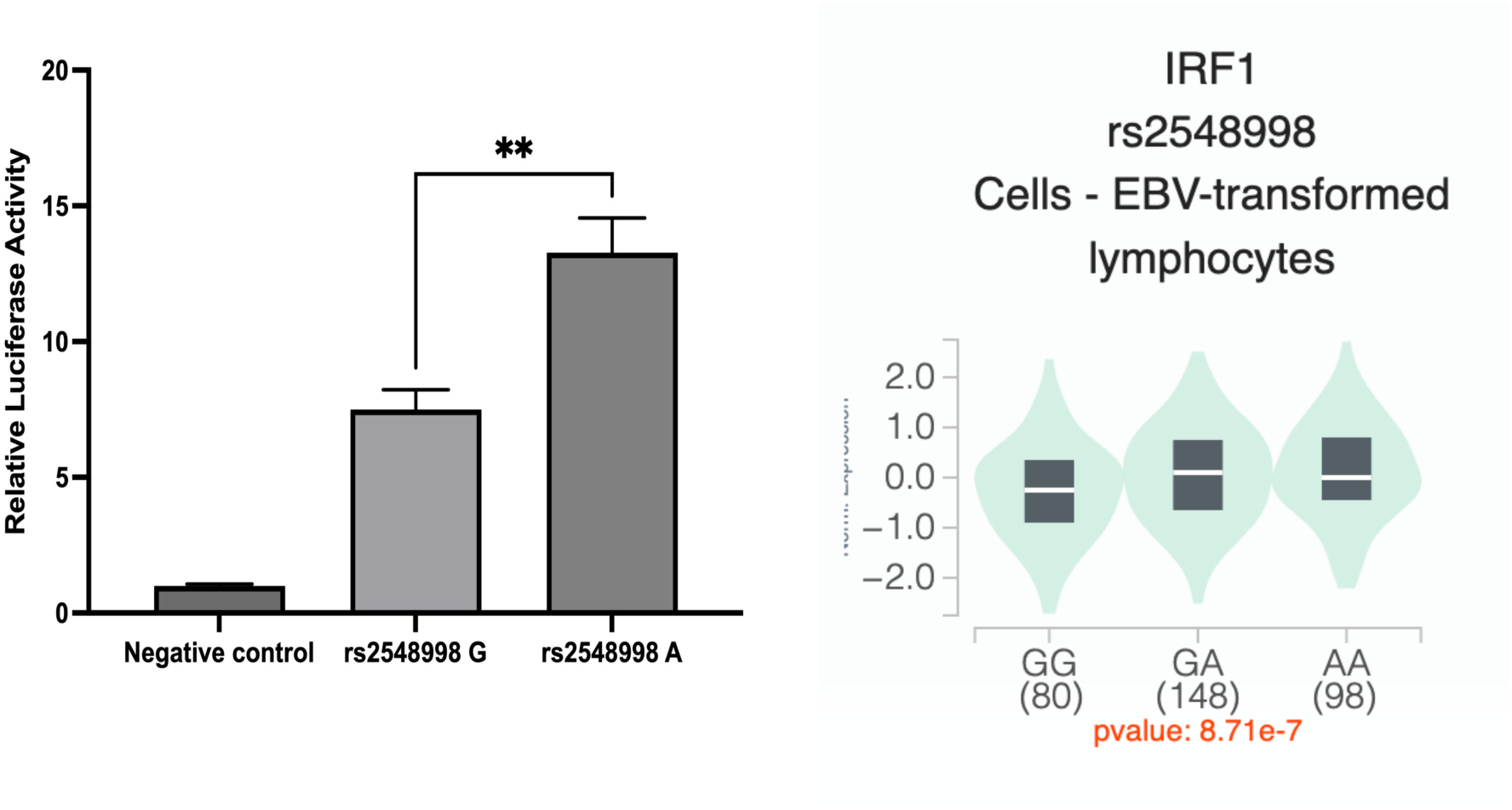
Left panel: Results of an enhancer reporter assay performed in CD3/CD28/CD2-activated primary human CD4+ T cells. We compared the G and A alleles of the SNP rs2548998, situated within the IRF1 promoter. The A allele increased reporter activity significantly when compared to the G allele. The negative control (negative control empty vector) is the basic pGL4.23 vector (which contains the SV40 promoter but is inefficient at driving luciferase expression). Data are represented as mean ± SEM of 4 biological replicates, and p values are calculated using the unpaired t-test. ** p<0.01. Right panel: violin plots derived from GTEx data in EBV-transformed lymphocytes. Human data corroborate the reporter assay, demonstrating that individuals carrying the A allele have higher expression of IRF1 that individuals carrying the G allele.

## Discussion

The field of genetics as applied to complex traits has started to move beyond the identification of genetic associations and toward the elucidation of the mechanisms through which genetic variants confer risk^31,32^. However, a significant impediment to accomplishing this task is the fact that the strength GWAS studies, which leverage LD to identify regions conferring genetic risk, is also a weakness, in that the SNPs that tag genetic risk loci are in LD with dozens, sometimes hundreds, of other SNPs, most of which have no influence at all on disease risk. Thus, distinguishing the true causal variants (i.e., those that exert the relevant biological effects) from the innocuous ones in which they are in LD, has been a challenge. At the same time, the discovery that, for most complex traits ^14^, including autoimmune diseases ^15^, genetic risk is likely to impinge on regulatory functions rather than the protein-coding sequences of pathology-driving genes, has complicated the search for target genes (i.e., the genes influenced by the causal variants).

In this study, we used chromatin features and the presence of PROseq signal, analyzed using dREG to identify CREs, to refine our understanding of the functional, non-coding regions on JIA risk haplotypes and thus narrow the search for causal variants. We corroborate our earlier papers^11,12^ showing that the JIA risk haplotypes are highly enriched (compared to randomly selected regions of the genome) for enhancers. We also demonstrate that the target genes, i.e., the genes most likely influenced by genetic variants within these functional regions, can be identified using high-resolution chromatin data (MicroC). The 3D chromatin data are essential to interpreting GWAS signal, as risk-conferring variants may not influence the “nearest gene” to the SNPs used to tag the locus^33^. Indeed, as Pelikan et al have shown, target genes for autoimmune diseases may not even be on the risk haplotypes^34^.

As expected, there were small differences in the SNPs and target genes that emerged from the analyses of activated vs resting CD4+ T cells, although most of the target genes identified in resting CD4+ T cells were also identified in activated cells (**Figure 1**). Previous *in vitro* modeling studies that showed that the effects of genetic variants on immune cell function may be context-dependent ^29,35^ as CD4+ T cells undergo chromatin remodeling during activation ^36^. However, we have published studies using HiChIP demonstrating that the 3D chromatin architecture of the CD4+ T cells from the peripheral blood of children with JIA largely resembles that of CD4+ T cells from healthy children ^37^. Note also that we demonstrated small differences in the candidate genes identified using non-specific activation with PMA compared with the genes identified after activating CD4+ T cells with CD3-CD28-IL2. These differences may simply reflect the different time frames over which the cells were incubated prior to performance of the PROseq and MicroC assays (2 hr for PMA and 5 days for CD3-CD28-IL2).

It’s useful to note that the most notable differences in target gene identification were in the comparisons between primary CD4+ T cells and (resting) Jurkat T cells. While there were multiple genes identified in all cell types, there were n=14 genes/transcripts that were identified in only in Jurkat. This suggests caution must be taken in interpreting results of studies aiming to identify genetic mechanisms of autoimmunity performed in Jurkat cells, as Ray et al have recently stated ^38^.

The genes identified on these analyses corroborate other studies investigating the genetics and pathobiology of JIA. For example, the identification of *IRF1* as a likely target gene mediating JIA genetic risk is consistent with the enhanced IFNγ signaling previously observed by Throm and colleagues in JIA CD4+ T cells and monocytes^18^. Furthermore, these authors observed downstream effects of IFNγ on the expression of *SOCS1*, a gene identified in our analysis of CD3-CD28-IL2-activated CD4+ cells. ERAP2 is an endoplasmic reticulum-resident aminopeptidase that plays an important role in antigen processing ^39^. Genetic polymorphisms in this region associated with multiple rheumatic diseases, and our findings corroborate a recent study demonstrating that SNPs mediating risk at this locus influence cis-regulatory elements that influence *ERAP2* expression ^40^.

This study also refines that of our previously published work ^13^ and that of Pudjihartono and colleagues ^41^ by linking specific functional elements within JIA risk regions to promoters rather than using HiC data to identify all potential interactions between SNPs on JIA risk haplotypes and target genes. Thus we have reduced the list of candidate target genes in CD4+ T cells from >200 (as identified in our earlier paper ^13^ and that of Pudjihartono et al ^41^) to n=41. We believe that the combined used of 3D chromatin data plus identifying functional regions on the JIA risk haplotypes (in this paper, using RNApol II localization-PROseq) provides a more precise genomic localization of where causal JIA alleles are likely to be operating.

Several interesting questions emerge from this study. On several of the JIA risk haplotypes, for example, we identified SNPs within more than one cis-regulatory element. For example, within the *IRF1* locus, we identified SNPs within both the *IRF1* promoter and an intergenic enhancer located upstream of the *IRF1* transcription start site (see Figure 6). SNPs within each of these regions were associated with allele-specific effects on *IRF1* expression in GTEx data. This observation may simply reflect that the fact that the true expression-influencing SNP is in strong LD with the others. However, this finding it also raises the possibility of there being more than one risk-driving SNP, each located in different cis-regulatory element and co-inherited, on JIA risk haplotypes. This possibility is being increasingly recognized for complex traits^42,43^, Our recently-published MPRA data^30^ are consistent with that interpretation. In that paper, we identified *multiple* SNPs within several JIA risk regions (including *IRF1* and *LNPEP-ERAP2*) that displayed intrinsic, allele-specific effects on gene expression, independent of LD. This question will need to be addressed, as it has implications for how we use genetic data to inform patient care. The availability targeted long genome sequencing^44^ should facilitate the identification of patterns of coinheritance that will allow us to determine the extent to which alleles identified by their intrinsic properties in functional assays are co-inherited.

### Conclusions

We demonstrate that, by a functional read-out of CREs (PROseq-PROcap) and high-resolution 3D chromatin mapping (MicroC), we can nominate a list of plausible causal variants on JIA risk haplotypes and their likely target genes. Findings from this study are corroborated by GTEx consortium data demonstrating allele-specific effects on gene expression in individuals carrying the nominated variants.

## Acknowledgements

This work was supported by R01 AR078785 and R21 AR076948 from the National Institutes of Health (JNJ) and by a summer medical student preceptorship (to EKH) from the Rheumatology Research Foundation.

## Data Availability

MicroC data is available on dbGAP, accession # phs002146.v1.p1. PROseq data is available on the Gene Expression Omnibus, accession # GSE85337.

