## Supplemental Tables for "Functional and Computational Interrogation of the Juvenile Idiopathic Arthritis Risk Loci Identifies Candidate Causal SNPs and Target Genes in CD4+ T cells"

**Supplemental Table 1: Chromosome coordinates for PROseq peaks in resting CD4+ T cells**

| chromosome | start | end | length | haplotype |
|---|---|---|---|---|
| chr1 | 64956710 | 64957090 | 380 | JAK1 |
| chr1 | 64962410 | 64962770 | 360 | JAK1 |
| chr1 | 64965120 | 64965370 | 250 | JAK1 |
| chr1 | 64965760 | 64966280 | 520 | JAK1 |
| chr1 | 64966300 | 64966770 | 470 | JAK1 |
| chr1 | 64966790 | 64967050 | 260 | JAK1 |
| chr1 | 113780730 | 113781150 | 420 | PTPN22 |
| chr1 | 113783450 | 113783800 | 350 | PTPN22 |
| chr1 | 113811490 | 113811920 | 430 | PTPN22 |
| chr1 | 113811940 | 113812130 | 190 | PTPN22 |
| chr1 | 113812150 | 113813110 | 960 | PTPN22 |
| chr1 | 153261480 | 153262000 | 520 | PRR9/LOR |
| chr1 | 154324660 | 154325120 | 460 | ATP8B2/IL6R |
| chr1 | 154325140 | 154325270 | 130 | ATP8B2/IL6R |
| chr1 | 154325290 | 154325640 | 350 | ATP8B2/IL6R |
| chr1 | 154325660 | 154325900 | 240 | ATP8B2/IL6R |
| chr1 | 154327480 | 154328070 | 590 | ATP8B2/IL6R |
| chr1 | 154328220 | 154328840 | 620 | ATP8B2/IL6R |
| chr1 | 154330740 | 154331030 | 290 | ATP8B2/IL6R |
| chr1 | 154334960 | 154335380 | 420 | ATP8B2/IL6R |
| chr1 | 154337040 | 154337910 | 870 | ATP8B2/IL6R |
| chr1 | 154342670 | 154342890 | 220 | ATP8B2/IL6R |
| chr1 | 154342910 | 154343470 | 560 | ATP8B2/IL6R |
| chr1 | 154345180 | 154345770 | 590 | ATP8B2/IL6R |
| chr1 | 154352610 | 154353100 | 490 | ATP8B2/IL6R |
| chr1 | 154353120 | 154353600 | 480 | ATP8B2/IL6R |
| chr1 | 154385440 | 154385860 | 420 | ATP8B2/IL6R |
| chr1 | 154385880 | 154386110 | 230 | ATP8B2/IL6R |
| chr1 | 154386130 | 154386240 | 110 | ATP8B2/IL6R |
| chr1 | 154386260 | 154387020 | 760 | ATP8B2/IL6R |
| chr1 | 154387040 | 154387360 | 320 | ATP8B2/IL6R |
| chr1 | 154398370 | 154398680 | 310 | ATP8B2/IL6R |
| chr1 | 154403160 | 154403570 | 410 | ATP8B2/IL6R |
| chr1 | 154404740 | 154405440 | 700 | ATP8B2/IL6R |
| chr1 | 154405460 | 154405740 | 280 | ATP8B2/IL6R |
| chr10 | 6036900 | 6037480 | 580 | IL2RA |
| chr10 | 6037840 | 6038410 | 570 | IL2RA |

| | | | | |
|---|---|---|---|---|
| chr10 | 6050100 | 6050320 | 220 | IL2RA |
| chr10 | 6050760 | 6051470 | 710 | IL2RA |
| chr10 | 6052520 | 6052910 | 390 | IL2RA |
| chr10 | 48804240 | 48804540 | 300 | WDFY4 |
| chr12 | 111404650 | 111404940 | 290 | SH2B3–ATXN2 |
| chr12 | 111405350 | 111406000 | 650 | SH2B3–ATXN2 |
| chr12 | 111406020 | 111406330 | 310 | SH2B3–ATXN2 |
| chr12 | 111406530 | 111407090 | 560 | SH2B3–ATXN2 |
| chr12 | 111434590 | 111434920 | 330 | SH2B3–ATXN2 |
| chr12 | 111434980 | 111435320 | 340 | SH2B3–ATXN2 |
| chr12 | 111598410 | 111598810 | 400 | SH2B3–ATXN2 |
| chr12 | 111598970 | 111599150 | 180 | SH2B3–ATXN2 |
| chr12 | 111599170 | 111599730 | 560 | SH2B3–ATXN2 |
| chr12 | 111599750 | 111600100 | 350 | SH2B3–ATXN2 |
| chr14 | 68788760 | 68789300 | 540 | ZFP36L1 |
| chr14 | 68789320 | 68789770 | 450 | ZFP36L1 |
| chr14 | 68790520 | 68791490 | 970 | ZFP36L1 |
| chr14 | 68792660 | 68792930 | 270 | ZFP36L1 |
| chr14 | 68792950 | 68793640 | 690 | ZFP36L1 |
| chr18 | 12775530 | 12775990 | 460 | PTPN2 |
| chr19 | 10333020 | 10333400 | 380 | TYK2 |
| chr19 | 10333420 | 10333810 | 390 | TYK2 |
| chr19 | 10333830 | 10334180 | 350 | TYK2 |
| chr19 | 10334200 | 10334560 | 360 | TYK2 |
| chr19 | 10338790 | 10339340 | 550 | TYK2 |
| chr19 | 10339360 | 10339510 | 150 | TYK2 |
| chr19 | 10339530 | 10340020 | 490 | TYK2 |
| chr19 | 10380430 | 10380950 | 520 | TYK2 |
| chr19 | 10380970 | 10381290 | 320 | TYK2 |
| chr21 | 35326470 | 35326840 | 370 | RUNX1 |
| chr22 | 21567380 | 21567830 | 450 | UBE2L3 |
| chr22 | 21567850 | 21568060 | 210 | UBE2L3 |
| chr22 | 21568080 | 21568680 | 600 | UBE2L3 |
| chr22 | 21568700 | 21568920 | 220 | UBE2L3 |
| chr22 | 30289470 | 30290210 | 740 | RNF215 |
| chr22 | 30290230 | 30290670 | 440 | RNF215 |
| chr22 | 30305230 | 30305890 | 660 | RNF215 |
| chr22 | 30305910 | 30306170 | 260 | RNF215 |
| chr22 | 30306190 | 30306460 | 270 | RNF215 |

| | | | | |
|---|---|---|---|---|
| chr22 | 30306480 | 30306730 | 250 | RNF215 |
| chr22 | 30306750 | 30307100 | 350 | RNF215 |
| chr22 | 30309840 | 30310220 | 380 | RNF215 |
| chr22 | 30324140 | 30324800 | 660 | RNF215 |
| chr22 | 30326800 | 30327630 | 830 | RNF215 |
| chr22 | 30356180 | 30356760 | 580 | RNF215 |
| chr22 | 30356780 | 30357180 | 400 | RNF215 |
| chr22 | 30362020 | 30362490 | 470 | RNF215 |
| chr22 | 30387300 | 30387840 | 540 | RNF215 |
| chr22 | 30396740 | 30397070 | 330 | RNF215 |
| chr22 | 30397220 | 30397550 | 330 | RNF215 |
| chr22 | 30397760 | 30398200 | 440 | RNF215 |
| chr3 | 122055180 | 122055390 | 210 | ILDR1/CD86 |
| chr4 | 122151860 | 122152620 | 760 | IL2–IL21 |
| chr4 | 122152640 | 122152810 | 170 | IL2–IL21 |
| chr4 | 122152830 | 122153240 | 410 | IL2–IL21 |
| chr4 | 122458540 | 122459000 | 460 | IL2–IL21 |
| chr4 | 122578370 | 122578520 | 150 | IL2–IL21 |
| chr4 | 122578540 | 122578990 | 450 | IL2–IL21 |
| chr4 | 122619540 | 122620290 | 750 | IL2–IL21 |
| chr5 | 56142490 | 56143120 | 630 | ANKRD55 |
| chr5 | 96885860 | 96886500 | 640 | ERAP2–LNPEP |
| chr5 | 96896170 | 96896800 | 630 | ERAP2–LNPEP |
| chr5 | 96934100 | 96934310 | 210 | ERAP2–LNPEP |
| chr5 | 96934330 | 96934580 | 250 | ERAP2–LNPEP |
| chr5 | 96934600 | 96935010 | 410 | ERAP2–LNPEP |
| chr5 | 96935030 | 96935480 | 450 | ERAP2–LNPEP |
| chr5 | 96935500 | 96935720 | 220 | ERAP2–LNPEP |
| chr5 | 96935740 | 96936190 | 450 | ERAP2–LNPEP |
| chr5 | 96936210 | 96936390 | 180 | ERAP2–LNPEP |
| chr5 | 96936710 | 96937140 | 430 | ERAP2–LNPEP |
| chr5 | 96962490 | 96962800 | 310 | ERAP2–LNPEP |
| chr5 | 96962820 | 96963250 | 430 | ERAP2–LNPEP |
| chr5 | 96973850 | 96974260 | 410 | ERAP2–LNPEP |
| chr5 | 132490000 | 132490310 | 310 | C5orf56/IRF1 |
| chr5 | 132490330 | 132490640 | 310 | C5orf56/IRF1 |
| chr5 | 132490660 | 132491170 | 510 | C5orf56/IRF1 |
| chr5 | 132491190 | 132491620 | 430 | C5orf56/IRF1 |
| chr5 | 132495590 | 132495940 | 350 | C5orf56/IRF1 |

| | | | | |
|---|---|---|---|---|
| chr5 | 132495960 | 132496680 | 720 | C5orf56/IRF1 |
| chr5 | 132496700 | 132497160 | 460 | C5orf56/IRF1 |
| chr7 | 107167860 | 107168270 | 410 | HBP1 |
| chr7 | 107168290 | 107168810 | 520 | HBP1 |
| chr7 | 107168830 | 107169110 | 280 | HBP1 |
| chr7 | 107169510 | 107170220 | 710 | HBP1 |

**Supplemental Table 2: PROseq peaks in CD3+ CD28+ IL2-activated CD4+ T cells**

| chromosome | start | end | length | haplotype |
|---|---|---|---|---|
| chr1 | 64956680 | 64957050 | 370 | JAK1 |
| chr1 | 64957070 | 64957210 | 140 | JAK1 |
| chr1 | 64962470 | 64962780 | 310 | JAK1 |
| chr1 | 64965700 | 64966250 | 550 | JAK1 |
| chr1 | 64966270 | 64966820 | 550 | JAK1 |
| chr1 | 64966840 | 64967000 | 160 | JAK1 |
| chr1 | 113780670 | 113781120 | 450 | PTPN22 |
| chr1 | 113783470 | 113784180 | 710 | PTPN22 |
| chr1 | 113784200 | 113784410 | 210 | PTPN22 |
| chr1 | 113811720 | 113812110 | 390 | PTPN22 |
| chr1 | 113812130 | 113812740 | 610 | PTPN22 |
| chr1 | 113812760 | 113813220 | 460 | PTPN22 |
| chr1 | 113813360 | 113813820 | 460 | PTPN22 |
| chr1 | 154324560 | 154325100 | 540 | ATP8B2/IL6R |
| chr1 | 154325120 | 154325170 | 50 | ATP8B2/IL6R |
| chr1 | 154325190 | 154325650 | 460 | ATP8B2/IL6R |
| chr1 | 154327420 | 154327800 | 380 | ATP8B2/IL6R |
| chr1 | 154328530 | 154328910 | 380 | ATP8B2/IL6R |
| chr1 | 154330760 | 154331100 | 340 | ATP8B2/IL6R |
| chr1 | 154334910 | 154335370 | 460 | ATP8B2/IL6R |
| chr1 | 154337210 | 154337920 | 710 | ATP8B2/IL6R |
| chr1 | 154342460 | 154342830 | 370 | ATP8B2/IL6R |
| chr1 | 154342850 | 154343350 | 500 | ATP8B2/IL6R |
| chr1 | 154353120 | 154353580 | 460 | ATP8B2/IL6R |
| chr1 | 154353600 | 154354060 | 460 | ATP8B2/IL6R |
| chr1 | 154385490 | 154385850 | 360 | ATP8B2/IL6R |
| chr1 | 154385870 | 154386070 | 200 | ATP8B2/IL6R |
| chr1 | 154386360 | 154386990 | 630 | ATP8B2/IL6R |
| chr1 | 154398070 | 154398660 | 590 | ATP8B2/IL6R |
| chr1 | 154403120 | 154403450 | 330 | ATP8B2/IL6R |
| chr1 | 154404940 | 154405480 | 540 | ATP8B2/IL6R |
| chr10 | 6045000 | 6045460 | 460 | IL2RA |
| chr10 | 6045920 | 6046440 | 520 | IL2RA |
| chr10 | 6050710 | 6051410 | 700 | IL2RA |
| chr10 | 6052600 | 6053180 | 580 | IL2RA |
| chr10 | 48800310 | 48800760 | 450 | WDFY4 |

| | | | | |
|---|---|---|---|---|
| chr12 | 111404550 | 111404790 | 240 | SH2B3–ATXN2 |
| chr12 | 111405270 | 111405610 | 340 | SH2B3–ATXN2 |
| chr12 | 111405630 | 111405980 | 350 | SH2B3–ATXN2 |
| chr12 | 111406000 | 111406230 | 230 | SH2B3–ATXN2 |
| chr12 | 111406250 | 111406500 | 250 | SH2B3–ATXN2 |
| chr12 | 111406520 | 111406760 | 240 | SH2B3–ATXN2 |
| chr12 | 111406780 | 111407040 | 260 | SH2B3–ATXN2 |
| chr12 | 111597000 | 111597670 | 670 | SH2B3–ATXN2 |
| chr12 | 111598850 | 111599160 | 310 | SH2B3–ATXN2 |
| chr12 | 111599180 | 111600150 | 970 | SH2B3–ATXN2 |
| chr14 | 68786770 | 68787160 | 390 | ZFP36L1 |
| chr14 | 68787180 | 68787550 | 370 | ZFP36L1 |
| chr14 | 68789000 | 68789240 | 240 | ZFP36L1 |
| chr14 | 68792130 | 68792410 | 280 | ZFP36L1 |
| chr14 | 68792750 | 68792940 | 190 | ZFP36L1 |
| chr14 | 68792960 | 68793650 | 690 | ZFP36L1 |
| chr18 | 12774880 | 12775330 | 450 | PTPN2 |
| chr18 | 12775350 | 12775760 | 410 | PTPN2 |
| chr18 | 12775780 | 12776020 | 240 | PTPN2 |
| chr18 | 12776040 | 12776390 | 350 | PTPN2 |
| chr19 | 10332830 | 10333390 | 560 | TYK2 |
| chr19 | 10333410 | 10334040 | 630 | TYK2 |
| chr19 | 10334060 | 10334360 | 300 | TYK2 |
| chr19 | 10339020 | 10339510 | 490 | TYK2 |
| chr19 | 10339530 | 10339820 | 290 | TYK2 |
| chr19 | 10339840 | 10340110 | 270 | TYK2 |
| chr19 | 10380370 | 10380930 | 560 | TYK2 |
| chr19 | 10380950 | 10381110 | 160 | TYK2 |
| chr21 | 35342370 | 35342800 | 430 | RUNX1 |
| chr21 | 35343160 | 35343460 | 300 | RUNX1 |
| chr21 | 35343480 | 35343910 | 430 | RUNX1 |
| chr22 | 21567480 | 21568070 | 590 | UBE2L3 |
| chr22 | 21568090 | 21568210 | 120 | UBE2L3 |
| chr22 | 21568230 | 21568560 | 330 | UBE2L3 |
| chr22 | 21568580 | 21569010 | 430 | UBE2L3 |
| chr22 | 21627200 | 21627520 | 320 | UBE2L3 |
| chr22 | 21628920 | 21629140 | 220 | UBE2L3 |
| chr22 | 30302250 | 30302760 | 510 | RNF215 |
| chr22 | 30305280 | 30305890 | 610 | RNF215 |

| | | | | |
|---|---|---|---|---|
| chr22 | 30305910 | 30306270 | 360 | RNF215 |
| chr22 | 30306290 | 30306770 | 480 | RNF215 |
| chr22 | 30307050 | 30307330 | 280 | RNF215 |
| chr22 | 30309880 | 30310160 | 280 | RNF215 |
| chr22 | 30324290 | 30324660 | 370 | RNF215 |
| chr22 | 30326760 | 30327280 | 520 | RNF215 |
| chr22 | 30327300 | 30327610 | 310 | RNF215 |
| chr22 | 30356320 | 30356750 | 430 | RNF215 |
| chr22 | 30356770 | 30357210 | 440 | RNF215 |
| chr22 | 30386850 | 30387270 | 420 | RNF215 |
| chr22 | 30387290 | 30387660 | 370 | RNF215 |
| chr22 | 30392630 | 30392930 | 300 | RNF215 |
| chr22 | 30396680 | 30397110 | 430 | RNF215 |
| chr3 | 46926800 | 46927420 | 620 | PTH1R |
| chr3 | 122025360 | 122025700 | 340 | ILDR1/CD86 |
| chr3 | 122030210 | 122031100 | 890 | ILDR1/CD86 |
| chr3 | 122066990 | 122067500 | 510 | ILDR1/CD86 |
| chr3 | 122067690 | 122068270 | 580 | ILDR1/CD86 |
| chr3 | 122074580 | 122074850 | 270 | ILDR1/CD86 |
| chr3 | 122074870 | 122075510 | 640 | ILDR1/CD86 |
| chr3 | 122076990 | 122077760 | 770 | ILDR1/CD86 |
| chr3 | 122081570 | 122081960 | 390 | ILDR1/CD86 |
| chr3 | 122082180 | 122082640 | 460 | ILDR1/CD86 |
| chr3 | 122084620 | 122085310 | 690 | ILDR1/CD86 |
| chr3 | 122092400 | 122092860 | 460 | ILDR1/CD86 |
| chr3 | 122094050 | 122094480 | 430 | ILDR1/CD86 |
| chr3 | 122094930 | 122095250 | 320 | ILDR1/CD86 |
| chr3 | 122097910 | 122098220 | 310 | ILDR1/CD86 |
| chr3 | 122100880 | 122101520 | 640 | ILDR1/CD86 |
| chr3 | 122101540 | 122101970 | 430 | ILDR1/CD86 |
| chr4 | 122151880 | 122152080 | 200 | IL2–IL21 |
| chr4 | 122152100 | 122152630 | 530 | IL2–IL21 |
| chr4 | 122152850 | 122153320 | 470 | IL2–IL21 |
| chr4 | 122455030 | 122455530 | 500 | IL2–IL21 |
| chr4 | 122456260 | 122456700 | 440 | IL2–IL21 |
| chr4 | 122456820 | 122457040 | 220 | IL2–IL21 |
| chr4 | 122458460 | 122459070 | 610 | IL2–IL21 |
| chr4 | 122542150 | 122542830 | 680 | IL2–IL21 |
| chr4 | 122542850 | 122543210 | 360 | IL2–IL21 |

| | | | | |
|---|---|---|---|---|
| chr4 | 122548320 | 122548960 | 640 | IL2–IL21 |
| chr4 | 122578030 | 122578880 | 850 | IL2–IL21 |
| chr4 | 122578900 | 122579410 | 510 | IL2–IL21 |
| chr4 | 122584460 | 122584870 | 410 | IL2–IL21 |
| chr4 | 122616780 | 122617420 | 640 | IL2–IL21 |
| chr5 | 56142480 | 56143080 | 600 | ANKRD55 |
| chr5 | 96885790 | 96886420 | 630 | ERAP2–LNPEP |
| chr5 | 96933000 | 96934070 | 1070 | ERAP2–LNPEP |
| chr5 | 96934310 | 96934620 | 310 | ERAP2–LNPEP |
| chr5 | 96934640 | 96934950 | 310 | ERAP2–LNPEP |
| chr5 | 96934970 | 96935460 | 490 | ERAP2–LNPEP |
| chr5 | 96935480 | 96935690 | 210 | ERAP2–LNPEP |
| chr5 | 96935710 | 96936330 | 620 | ERAP2–LNPEP |
| chr5 | 96936660 | 96937150 | 490 | ERAP2–LNPEP |
| chr5 | 96960950 | 96961530 | 580 | ERAP2–LNPEP |
| chr5 | 96962500 | 96963150 | 650 | ERAP2–LNPEP |
| chr5 | 96963170 | 96963770 | 600 | ERAP2–LNPEP |
| chr5 | 132481160 | 132481360 | 200 | C5orf56/IRF1 |
| chr5 | 132488030 | 132488370 | 340 | C5orf56/IRF1 |
| chr5 | 132488630 | 132488960 | 330 | C5orf56/IRF1 |
| chr5 | 132490420 | 132490620 | 200 | C5orf56/IRF1 |
| chr5 | 132490640 | 132491390 | 750 | C5orf56/IRF1 |
| chr5 | 132495570 | 132496130 | 560 | C5orf56/IRF1 |
| chr5 | 132496150 | 132496770 | 620 | C5orf56/IRF1 |
| chr5 | 132496790 | 132497340 | 550 | C5orf56/IRF1 |
| chr6 | 135322500 | 135322900 | 400 | AHI1/LINC00271 |
| chr6 | 135330400 | 135330740 | 340 | AHI1/LINC00271 |
| chr7 | 107167600 | 107167860 | 260 | HBP1 |
| chr7 | 107167880 | 107168210 | 330 | HBP1 |
| chr7 | 107168230 | 107168440 | 210 | HBP1 |
| chr7 | 107168460 | 107168780 | 320 | HBP1 |
| chr7 | 107168800 | 107169090 | 290 | HBP1 |
| chr7 | 107169320 | 107169600 | 280 | HBP1 |

**Supplemental Table 3: Chromosome coordinates for PROseq peaks in PMA-activated CD4+ T cells**

| chromosome | start | end | length | haplotype |
|---|---|---|---|---|
| 1 | 64962510 | 64962760 | 250 | JAK1 |
| 1 | 64965090 | 64965420 | 330 | JAK1 |
| 1 | 64965710 | 64966210 | 500 | JAK1 |
| 1 | 64966400 | 64966830 | 430 | JAK1 |
| 1 | 113783520 | 113783760 | 240 | PTPN22 |
| 1 | 113811700 | 113812140 | 440 | PTPN22 |
| 1 | 113812160 | 113813120 | 960 | PTPN22 |
| 1 | 113813140 | 113813510 | 370 | PTPN22 |
| 1 | 153261620 | 153262050 | 430 | PRR9/LOR |
| 1 | 154325300 | 154325650 | 350 | ATP8B2/IL6R |
| 1 | 154325670 | 154325980 | 310 | ATP8B2/IL6R |
| 1 | 154326790 | 154327110 | 320 | ATP8B2/IL6R |
| 1 | 154327500 | 154327760 | 260 | ATP8B2/IL6R |
| 1 | 154328480 | 154328770 | 290 | ATP8B2/IL6R |
| 1 | 154330750 | 154331000 | 250 | ATP8B2/IL6R |
| 1 | 154334900 | 154335440 | 540 | ATP8B2/IL6R |
| 1 | 154342850 | 154343290 | 440 | ATP8B2/IL6R |
| 1 | 154352580 | 154353030 | 450 | ATP8B2/IL6R |
| 1 | 154353050 | 154353520 | 470 | ATP8B2/IL6R |
| 1 | 154385530 | 154386000 | 470 | ATP8B2/IL6R |
| 1 | 154386470 | 154387100 | 630 | ATP8B2/IL6R |
| 1 | 154396580 | 154396860 | 280 | ATP8B2/IL6R |
| 1 | 154404700 | 154405580 | 880 | ATP8B2/IL6R |
| 1 | 154405930 | 154406620 | 690 | ATP8B2/IL6R |
| 10 | 6036820 | 6037680 | 860 | IL2RA |
| 10 | 6038030 | 6038610 | 580 | IL2RA |
| 10 | 6045050 | 6045470 | 420 | IL2RA |
| 10 | 6045660 | 6046030 | 370 | IL2RA |
| 10 | 6046050 | 6046440 | 390 | IL2RA |
| 10 | 6049770 | 6050260 | 490 | IL2RA |
| 10 | 6050870 | 6051270 | 400 | IL2RA |
| 10 | 6051290 | 6051650 | 360 | IL2RA |
| 10 | 6051670 | 6052190 | 520 | IL2RA |
| 10 | 6052520 | 6053000 | 480 | IL2RA |
| 12 | 111403260 | 111403590 | 330 | SH2B3–ATXN2 |
| 12 | 111404330 | 111405070 | 740 | SH2B3–ATXN2 |

| | | | | |
|---|---|---|---|---|
| 12 | 111405340 | 111405550 | 210 | SH2B3–ATXN2 |
| 12 | 111405570 | 111406010 | 440 | SH2B3–ATXN2 |
| 12 | 111406030 | 111406360 | 330 | SH2B3–ATXN2 |
| 12 | 111406380 | 111406710 | 330 | SH2B3–ATXN2 |
| 12 | 111406730 | 111407280 | 550 | SH2B3–ATXN2 |
| 12 | 111412250 | 111412670 | 420 | SH2B3–ATXN2 |
| 12 | 111430220 | 111430490 | 270 | SH2B3–ATXN2 |
| 12 | 111430680 | 111431200 | 520 | SH2B3–ATXN2 |
| 12 | 111434490 | 111434830 | 340 | SH2B3–ATXN2 |
| 12 | 111434850 | 111435460 | 610 | SH2B3–ATXN2 |
| 12 | 111546780 | 111547320 | 540 | SH2B3–ATXN2 |
| 12 | 111599160 | 111599740 | 580 | SH2B3–ATXN2 |
| 12 | 111599760 | 111600100 | 340 | SH2B3–ATXN2 |
| 12 | 111600120 | 111600360 | 240 | SH2B3–ATXN2 |
| 14 | 68785920 | 68786360 | 440 | ZFP36L1 |
| 14 | 68789110 | 68789710 | 600 | ZFP36L1 |
| 14 | 68789730 | 68790060 | 330 | ZFP36L1 |
| 14 | 68792590 | 68792990 | 400 | ZFP36L1 |
| 14 | 68793010 | 68793240 | 230 | ZFP36L1 |
| 14 | 68793260 | 68793780 | 520 | ZFP36L1 |
| 14 | 68794690 | 68795300 | 610 | ZFP36L1 |
| 19 | 10332970 | 10333400 | 430 | TYK2 |
| 19 | 10333420 | 10334090 | 670 | TYK2 |
| 19 | 10380470 | 10381120 | 650 | TYK2 |
| 19 | 10381140 | 10381470 | 330 | TYK2 |
| 22 | 21567490 | 21567840 | 350 | UBE2L3 |
| 22 | 21567860 | 21568100 | 240 | UBE2L3 |
| 22 | 21568120 | 21568520 | 400 | UBE2L3 |
| 22 | 21568540 | 21569020 | 480 | UBE2L3 |
| 22 | 30305360 | 30305750 | 390 | RNF215 |
| 22 | 30326330 | 30326730 | 400 | RNF215 |
| 22 | 30326830 | 30327470 | 640 | RNF215 |
| 22 | 30356230 | 30356750 | 520 | RNF215 |
| 22 | 30356770 | 30357370 | 600 | RNF215 |
| 22 | 30358060 | 30358370 | 310 | RNF215 |
| 22 | 30362220 | 30362510 | 290 | RNF215 |
| 22 | 30396630 | 30397090 | 460 | RNF215 |
| 22 | 30397110 | 30397530 | 420 | RNF215 |
| 22 | 30397550 | 30397760 | 210 | RNF215 |

| | | | | |
|---|---|---|---|---|
| 22 | 30397780 | 30398330 | 550 | RNF215 |
| 3 | 122055140 | 122055410 | 270 | ILDR1/CD86 |
| 4 | 122151810 | 122152130 | 320 | IL2–IL21 |
| 4 | 122152150 | 122152600 | 450 | IL2–IL21 |
| 4 | 122152620 | 122152870 | 250 | IL2–IL21 |
| 4 | 122453670 | 122454390 | 720 | IL2–IL21 |
| 4 | 122456180 | 122456550 | 370 | IL2–IL21 |
| 4 | 122456570 | 122457070 | 500 | IL2–IL21 |
| 4 | 122457660 | 122458240 | 580 | IL2–IL21 |
| 4 | 122458260 | 122459090 | 830 | IL2–IL21 |
| 4 | 122502960 | 122503780 | 820 | IL2–IL21 |
| 4 | 122504080 | 122504550 | 470 | IL2–IL21 |
| 4 | 122507530 | 122507820 | 290 | IL2–IL21 |
| 4 | 122507840 | 122508530 | 690 | IL2–IL21 |
| 4 | 122534250 | 122534640 | 390 | IL2–IL21 |
| 4 | 122539910 | 122540740 | 830 | IL2–IL21 |
| 4 | 122541370 | 122541900 | 530 | IL2–IL21 |
| 4 | 122542340 | 122542720 | 380 | IL2–IL21 |
| 4 | 122542890 | 122543160 | 270 | IL2–IL21 |
| 4 | 122578240 | 122578670 | 430 | IL2–IL21 |
| 4 | 122578690 | 122578890 | 200 | IL2–IL21 |
| 4 | 122578910 | 122579480 | 570 | IL2–IL21 |
| 4 | 122617620 | 122618120 | 500 | IL2–IL21 |
| 4 | 122619120 | 122619630 | 510 | IL2–IL21 |
| 5 | 56142730 | 56143300 | 570 | ANKRD55 |
| 5 | 96885900 | 96886470 | 570 | ERAP2–LNPEP |
| 5 | 96893290 | 96893740 | 450 | ERAP2–LNPEP |
| 5 | 96896200 | 96896660 | 460 | ERAP2–LNPEP |
| 5 | 96896920 | 96897310 | 390 | ERAP2–LNPEP |
| 5 | 96934110 | 96934570 | 460 | ERAP2–LNPEP |
| 5 | 96934590 | 96935010 | 420 | ERAP2–LNPEP |
| 5 | 96935030 | 96935480 | 450 | ERAP2–LNPEP |
| 5 | 96935500 | 96935700 | 200 | ERAP2–LNPEP |
| 5 | 96935720 | 96936180 | 460 | ERAP2–LNPEP |
| 5 | 96936610 | 96937010 | 400 | ERAP2–LNPEP |
| 5 | 132490320 | 132491310 | 990 | C5orf56/IRF1 |
| 5 | 132491330 | 132491960 | 630 | C5orf56/IRF1 |
| 5 | 132495560 | 132495970 | 410 | C5orf56/IRF1 |
| 5 | 132495990 | 132496200 | 210 | C5orf56/IRF1 |

| | | | | |
|---|---|---|---|---|
| 5 | 132496220 | 132497020 | 800 | C5orf56/IRF1 |
| 7 | 107167590 | 107168360 | 770 | HBP1 |
| 7 | 107168380 | 107168630 | 250 | HBP1 |
| 7 | 107168650 | 107169120 | 470 | HBP1 |
| 7 | 107169140 | 107169450 | 310 | HBP1 |
| 7 | 107169470 | 107170090 | 620 | HBP1 |
| 7 | 107170110 | 107170430 | 320 | HBP1 |

**Supplemental Table 4: JIA SNPs within PROseq peaks in resting CD4+ T cells**

| chromosome | start | end | SNP |
|---|---|---|---|
| 1 | 154328769 | 154328770 | rs1194611 |
| 1 | 154325897 | 154325898 | rs1205591 |
| 1 | 113758712 | 113758713 | rs61817589 |
| 1 | 154386702 | 154386703 | rs6427627 |
| 1 | 64966236 | 64966237 | rs72922282 |
| 1 | 64965275 | 64965276 | rs76454749 |
| 1 | 154386934 | 154386935 | rs9651053 |
| 2 | 100142822 | 100142823 | rs11123810 |
| 2 | 100142615 | 100142616 | rs13003982 |
| 2 | 100142567 | 100142568 | rs17023563 |
| 2 | 100140845 | 100140846 | rs4850920 |
| 2 | 100140817 | 100140818 | rs4851252 |
| 2 | 100142406 | 100142407 | rs77884588 |
| 3 | 46405229 | 46405230 | rs11574428 |
| 3 | 46405318 | 46405319 | rs11574429 |
| 3 | 46948413 | 46948414 | rs12491473 |
| 3 | 47163448 | 47163449 | rs13078131 |
| 3 | 46370816 | 46370817 | rs1800023 |
| 3 | 46371926 | 46371927 | rs2254089 |
| 3 | 46371842 | 46371843 | rs2856762 |
| 3 | 46372458 | 46372459 | rs2856765 |
| 3 | 46206893 | 46206894 | rs3176824 |
| 3 | 46206732 | 46206733 | rs3176825 |
| 3 | 46208437 | 46208438 | rs3181080 |
| 3 | 46208230 | 46208231 | rs34423195 |
| 3 | 46100971 | 46100972 | rs34460587 |
| 3 | 46208516 | 46208517 | rs34919616 |
| 3 | 46368645 | 46368646 | rs41490645 |
| 3 | 46372543 | 46372544 | rs41515644 |
| 3 | 46111008 | 46111009 | rs4682810 |
| 3 | 46098580 | 46098581 | rs71327024 |
| 3 | 46368544 | 46368545 | rs7637813 |
| 3 | 46948337 | 46948338 | rs7640397 |
| 5 | 132490629 | 132490630 | rs10900809 |
| 5 | 96885860 | 96885861 | rs11135483 |
| 5 | 96886184 | 96886185 | rs11135484 |
| 5 | 132490720 | 132490721 | rs11242115 |

| | | | |
|---|---|---|---|
| 5 | 96876036 | 96876037 | rs1230358 |
| 5 | 96877190 | 96877191 | rs1230360 |
| 5 | 96886315 | 96886316 | rs13167902 |
| 5 | 96963053 | 96963054 | rs1423357 |
| 5 | 96936802 | 96936803 | rs17684739 |
| 5 | 132490149 | 132490150 | rs2070721 |
| 5 | 96886478 | 96886479 | rs2303208 |
| 5 | 96886480 | 96886481 | rs2303209 |
| 5 | 96936715 | 96936716 | rs2548224 |
| 5 | 132496821 | 132496822 | rs2548998 |
| 5 | 132491498 | 132491499 | rs2549005 |
| 5 | 132491372 | 132491373 | rs2549006 |
| 5 | 132491072 | 132491073 | rs2549009 |
| 5 | 96881079 | 96881080 | rs2549780 |
| 5 | 96935933 | 96935934 | rs2617434 |
| 5 | 132496073 | 132496074 | rs2706385 |
| 5 | 132496994 | 132496995 | rs2706386 |
| 5 | 132490617 | 132490618 | rs960757 |
| 5 | 96877076 | 96877077 | rs968218 |
| 6 | 135497758 | 135497759 | rs13197384 |
| 7 | 28180608 | 28180609 | rs10255700 |
| 7 | 107168486 | 107168487 | rs112527862 |
| 7 | 22727025 | 22727026 | rs1800795 |
| 7 | 107168034 | 107168035 | rs257387 |
| 7 | 107168610 | 107168611 | rs4730220 |
| 7 | 107169847 | 107169848 | rs4730222 |
| 7 | 107169661 | 107169662 | rs61014091 |
| 7 | 107561129 | 107561130 | rs62483725 |
| 10 | 48804277 | 48804278 | rs10857649 |
| 10 | 6050129 | 6050130 | rs10905669 |
| 10 | 6072892 | 6072893 | rs10905718 |
| 10 | 86971004 | 86971005 | rs1240380 |
| 10 | 6072696 | 6072697 | rs41295061 |
| 10 | 6052733 | 6052734 | rs61839660 |
| 10 | 6069111 | 6069112 | rs7893324 |
| 12 | 6383933 | 6383934 | rs28999107 |
| 12 | 111599195 | 111599196 | rs695871 |
| 12 | 111599124 | 111599125 | rs695872 |
| 13 | 42396586 | 42396587 | rs12874234 |

| | | | |
|---|---|---|---|
| 13 | 42396676 | 42396677 | rs12874725 |
| 13 | 42393090 | 42393091 | rs34264781 |
| 13 | 42393136 | 42393137 | rs34700891 |
| 13 | 39655753 | 39655754 | rs3812882 |
| 13 | 39655819 | 39655820 | rs3812883 |
| 13 | 39655905 | 39655906 | rs3812884 |
| 13 | 39655984 | 39655985 | rs3812885 |
| 13 | 39656078 | 39656079 | rs3812886 |
| 13 | 39656091 | 39656092 | rs3812887 |
| 13 | 39656156 | 39656157 | rs3812888 |
| 13 | 42405814 | 42405815 | rs7326472 |
| 13 | 39655586 | 39655587 | rs7327779 |
| 13 | 42392749 | 42392750 | rs7987414 |
| 13 | 42392875 | 42392876 | rs7987728 |
| 13 | 39655230 | 39655231 | rs9548865 |
| 13 | 39655282 | 39655283 | rs9548866 |
| 13 | 39655430 | 39655431 | rs9548867 |
| 13 | 39655606 | 39655607 | rs9603589 |
| 14 | 68793572 | 68793573 | rs10443 |
| 14 | 68780587 | 68780588 | rs11624323 |
| 14 | 68792688 | 68792689 | rs11847049 |
| 14 | 68792784 | 68792785 | rs11851414 |
| 14 | 68820765 | 68820766 | rs194742 |
| 14 | 68791140 | 68791141 | rs36045050 |
| 14 | 68780762 | 68780763 | rs56119720 |
| 14 | 68781230 | 68781231 | rs56232028 |
| 14 | 68793310 | 68793311 | rs57786342 |
| 14 | 68781539 | 68781540 | rs72731550 |
| 14 | 68782137 | 68782138 | rs72731551 |
| 14 | 68820939 | 68820940 | rs72731567 |
| 14 | 68780268 | 68780269 | rs76200426 |
| 16 | 11312945 | 11312946 | rs7191700 |
| 16 | 11312934 | 11312935 | rs72773819 |
| 18 | 12775851 | 12775852 | rs2542147 |
| 18 | 12775963 | 12775964 | rs2847258 |
| 18 | 12775821 | 12775822 | rs2847259 |
| 18 | 12775591 | 12775592 | rs2847260 |
| 19 | 10315347 | 10315348 | rs1124765 |
| 19 | 10334555 | 10334556 | rs116900565 |

| | | | |
|---|---|---|---|
| 19 | 10295754 | 10295755 | rs12972990 |
| 19 | 10334149 | 10334150 | rs2278442 |
| 19 | 10339608 | 10339609 | rs281414 |
| 19 | 10315309 | 10315310 | rs368835 |
| 19 | 10315745 | 10315746 | rs378395 |
| 22 | 30423639 | 30423640 | rs1061660 |
| 22 | 30423769 | 30423770 | rs1061664 |
| 22 | 21568166 | 21568167 | rs11089620 |
| 22 | 21630278 | 21630279 | rs11704601 |
| 22 | 30356952 | 30356953 | rs13053375 |
| 22 | 21567396 | 21567397 | rs140490 |
| 22 | 21568854 | 21568855 | rs140492 |
| 22 | 30356351 | 30356352 | rs17657653 |
| 22 | 30424088 | 30424089 | rs17658686 |
| 22 | 30426472 | 30426473 | rs2072158 |
| 22 | 21568614 | 21568615 | rs2266959 |
| 22 | 21630089 | 21630090 | rs3747093 |
| 22 | 30290236 | 30290237 | rs4823085 |
| 22 | 30387535 | 30387536 | rs5753116 |
| 22 | 30387561 | 30387562 | rs5753117 |
| 22 | 30356706 | 30356707 | rs5994293 |
| 22 | 30396924 | 30396925 | rs740084 |
| 22 | 30397280 | 30397281 | rs887098 |

**Supplemental Table 5: JIA SNPs within PROseq peaks in CD3+ CD28+ IL2-activated CD4+ T cells**

| chromosome | start | end | SNP |
|---|---|---|---|
| chr1 | 154328769 | 154328770 | rs1194611 |
| chr1 | 113813502 | 113813503 | rs1217411 |
| chr1 | 113758712 | 113758713 | rs61817589 |
| chr1 | 154386702 | 154386703 | rs6427627 |
| chr1 | 64966236 | 64966237 | rs72922282 |
| chr1 | 154386934 | 154386935 | rs9651053 |
| chr2 | 100142822 | 100142823 | rs11123810 |
| chr2 | 100142615 | 100142616 | rs13003982 |
| chr2 | 100142567 | 100142568 | rs17023563 |
| chr2 | 100140845 | 100140846 | rs4850920 |
| chr2 | 100140817 | 100140818 | rs4851252 |
| chr3 | 46953751 | 46953752 | rs10461018 |
| chr3 | 122074660 | 122074661 | rs10511408 |
| chr3 | 122077241 | 122077242 | rs11922347 |
| chr3 | 46948413 | 46948414 | rs12491473 |
| chr3 | 46370816 | 46370817 | rs1800023 |
| chr3 | 46369444 | 46369445 | rs2734225 |
| chr3 | 46370169 | 46370170 | rs2856758 |
| chr3 | 46372458 | 46372459 | rs2856765 |
| chr3 | 46208437 | 46208438 | rs3181080 |
| chr3 | 46208230 | 46208231 | rs34423195 |
| chr3 | 46208516 | 46208517 | rs34919616 |
| chr3 | 122077384 | 122077385 | rs36226551 |
| chr3 | 46368645 | 46368646 | rs41490645 |
| chr3 | 46372543 | 46372544 | rs41515644 |
| chr3 | 46097803 | 46097804 | rs62243876 |
| chr3 | 46098580 | 46098581 | rs71327024 |
| chr3 | 46368544 | 46368545 | rs7637813 |
| chr3 | 46948337 | 46948338 | rs7640397 |
| chr5 | 132488234 | 132488235 | rs10035166 |
| chr5 | 132481210 | 132481211 | rs10072700 |
| chr5 | 96885860 | 96885861 | rs11135483 |
| chr5 | 96886184 | 96886185 | rs11135484 |
| chr5 | 132490720 | 132490721 | rs11242115 |
| chr5 | 96876036 | 96876037 | rs1230358 |
| chr5 | 96877190 | 96877191 | rs1230360 |

| | | | |
|---|---|---|---|
| chr5 | 96877466 | 96877467 | rs1230361 |
| chr5 | 96886315 | 96886316 | rs13167902 |
| chr5 | 96963053 | 96963054 | rs1423357 |
| chr5 | 96933702 | 96933703 | rs17487596 |
| chr5 | 96936802 | 96936803 | rs17684739 |
| chr5 | 96933157 | 96933158 | rs1981846 |
| chr5 | 132488793 | 132488794 | rs2070722 |
| chr5 | 96936715 | 96936716 | rs2548224 |
| chr5 | 132496821 | 132496822 | rs2548998 |
| chr5 | 132491372 | 132491373 | rs2549006 |
| chr5 | 132491182 | 132491183 | rs2549007 |
| chr5 | 132491072 | 132491073 | rs2549009 |
| chr5 | 96935933 | 96935934 | rs2617434 |
| chr5 | 132491187 | 132491188 | rs2706384 |
| chr5 | 132496073 | 132496074 | rs2706385 |
| chr5 | 132496994 | 132496995 | rs2706386 |
| chr5 | 132490617 | 132490618 | rs960757 |
| chr5 | 96877076 | 96877077 | rs968218 |
| chr6 | 135497758 | 135497759 | rs13197384 |
| chr7 | 107168486 | 107168487 | rs112527862 |
| chr7 | 22723389 | 22723390 | rs12700386 |
| chr7 | 22726601 | 22726602 | rs1800797 |
| chr7 | 107168034 | 107168035 | rs257387 |
| chr7 | 107168610 | 107168611 | rs4730220 |
| chr7 | 107169847 | 107169848 | rs4730222 |
| chr7 | 107169661 | 107169662 | rs61014091 |
| chr7 | 107169438 | 107169439 | rs62483623 |
| chr10 | 6072892 | 6072893 | rs10905718 |
| chr10 | 86971004 | 86971005 | rs1240380 |
| chr10 | 6072696 | 6072697 | rs41295061 |
| chr10 | 6052733 | 6052734 | rs61839660 |
| chr10 | 6068865 | 6068866 | rs7090512 |
| chr10 | 6068911 | 6068912 | rs7090530 |
| chr10 | 6069111 | 6069112 | rs7893324 |
| chr12 | 111406413 | 111406414 | rs3742003 |
| chr12 | 111599195 | 111599196 | rs695871 |
| chr12 | 111599124 | 111599125 | rs695872 |
| chr13 | 42395509 | 42395510 | rs10507507 |
| chr13 | 42396586 | 42396587 | rs12874234 |

| | | | |
|---|---|---|---|
| chr13 | 42396676 | 42396677 | rs12874725 |
| chr13 | 42406741 | 42406742 | rs2324878 |
| chr13 | 42395369 | 42395370 | rs34182380 |
| chr13 | 42393136 | 42393137 | rs34700891 |
| chr13 | 39655753 | 39655754 | rs3812882 |
| chr13 | 39655819 | 39655820 | rs3812883 |
| chr13 | 39655905 | 39655906 | rs3812884 |
| chr13 | 39656078 | 39656079 | rs3812886 |
| chr13 | 39656091 | 39656092 | rs3812887 |
| chr13 | 39656156 | 39656157 | rs3812888 |
| chr13 | 42395062 | 42395063 | rs71428876 |
| chr13 | 42405814 | 42405815 | rs7326472 |
| chr13 | 39655586 | 39655587 | rs7327779 |
| chr13 | 42392749 | 42392750 | rs7987414 |
| chr13 | 42392875 | 42392876 | rs7987728 |
| chr13 | 42397077 | 42397078 | rs7989519 |
| chr13 | 39655282 | 39655283 | rs9548866 |
| chr13 | 39655430 | 39655431 | rs9548867 |
| chr13 | 39655606 | 39655607 | rs9603589 |
| chr14 | 68793572 | 68793573 | rs10443 |
| chr14 | 68787537 | 68787538 | rs11158764 |
| chr14 | 68780587 | 68780588 | rs11624323 |
| chr14 | 68792784 | 68792785 | rs11851414 |
| chr14 | 68822594 | 68822595 | rs1274955 |
| chr14 | 68820765 | 68820766 | rs194742 |
| chr14 | 68787266 | 68787267 | rs4899257 |
| chr14 | 68787473 | 68787474 | rs4902647 |
| chr14 | 68780762 | 68780763 | rs56119720 |
| chr14 | 68781230 | 68781231 | rs56232028 |
| chr14 | 68793310 | 68793311 | rs57786342 |
| chr14 | 68781539 | 68781540 | rs72731550 |
| chr14 | 68782137 | 68782138 | rs72731551 |
| chr14 | 68820939 | 68820940 | rs72731567 |
| chr14 | 68780268 | 68780269 | rs76200426 |
| chr14 | 68787353 | 68787354 | rs80192296 |
| chr16 | 11310999 | 11311000 | rs1345880 |
| chr16 | 11345781 | 11345782 | rs918737 |
| chr16 | 11345821 | 11345822 | rs918738 |
| chr16 | 11345875 | 11345876 | rs918739 |

| | | | |
|---|---|---|---|
| chr18 | 12775851 | 12775852 | rs2542147 |
| chr18 | 12776171 | 12776172 | rs2847256 |
| chr18 | 12776050 | 12776051 | rs2847257 |
| chr18 | 12775963 | 12775964 | rs2847258 |
| chr18 | 12775821 | 12775822 | rs2847259 |
| chr18 | 12775591 | 12775592 | rs2847260 |
| chr18 | 12775295 | 12775296 | rs2847262 |
| chr18 | 12774894 | 12774895 | rs888270 |
| chr18 | 12774981 | 12774982 | rs888271 |
| chr19 | 10315347 | 10315348 | rs1124765 |
| chr19 | 10295754 | 10295755 | rs12972990 |
| chr19 | 10334149 | 10334150 | rs2278442 |
| chr19 | 10289227 | 10289228 | rs2569693 |
| chr19 | 10339608 | 10339609 | rs281414 |
| chr19 | 10289433 | 10289434 | rs281439 |
| chr19 | 10289627 | 10289628 | rs281440 |
| chr19 | 10315309 | 10315310 | rs368835 |
| chr19 | 10315745 | 10315746 | rs378395 |
| chr19 | 10291454 | 10291455 | rs901886 |
| chr22 | 30423639 | 30423640 | rs1061660 |
| chr22 | 30423769 | 30423770 | rs1061664 |
| chr22 | 21568166 | 21568167 | rs11089620 |
| chr22 | 21630278 | 21630279 | rs11704601 |
| chr22 | 30356952 | 30356953 | rs13053375 |
| chr22 | 21568854 | 21568855 | rs140492 |
| chr22 | 30356351 | 30356352 | rs17657653 |
| chr22 | 30424088 | 30424089 | rs17658686 |
| chr22 | 30426472 | 30426473 | rs2072158 |
| chr22 | 21568614 | 21568615 | rs2266959 |
| chr22 | 21628970 | 21628971 | rs2298429 |
| chr22 | 21630089 | 21630090 | rs3747093 |
| chr22 | 30387535 | 30387536 | rs5753116 |
| chr22 | 30387561 | 30387562 | rs5753117 |
| chr22 | 30356706 | 30356707 | rs5994293 |
| chr22 | 21629915 | 21629916 | rs710177 |
| chr22 | 30396924 | 30396925 | rs740084 |

**Supplemental Table 6: JIA SNPs within PROseq peaks in PMA-activated CD4+ T cells**

| chromosome | start | end | SNP |
|---|---|---|---|
| 1 | 154328769 | 154328770 | rs1194611 |
| 1 | 154325897 | 154325898 | rs1205591 |
| 1 | 113813502 | 113813503 | rs1217411 |
| 1 | 154386702 | 154386703 | rs6427627 |
| 1 | 64965275 | 64965276 | rs76454749 |
| 1 | 154386934 | 154386935 | rs9651053 |
| 2 | 100142615 | 100142616 | rs13003982 |
| 2 | 100142567 | 100142568 | rs17023563 |
| 3 | 46959115 | 46959116 | rs11130110 |
| 3 | 46405229 | 46405230 | rs11574428 |
| 3 | 46405318 | 46405319 | rs11574429 |
| 3 | 46406423 | 46406424 | rs11574434 |
| 3 | 46406480 | 46406481 | rs11574435 |
| 3 | 46407899 | 46407900 | rs11574442 |
| 3 | 46948413 | 46948414 | rs12491473 |
| 3 | 47163448 | 47163449 | rs13078131 |
| 3 | 46109444 | 46109445 | rs1500004 |
| 3 | 46108907 | 46108908 | rs1546077 |
| 3 | 46370816 | 46370817 | rs1800023 |
| 3 | 46370169 | 46370170 | rs2856758 |
| 3 | 46208437 | 46208438 | rs3181080 |
| 3 | 46208230 | 46208231 | rs34423195 |
| 3 | 46100971 | 46100972 | rs34460587 |
| 3 | 46208516 | 46208517 | rs34919616 |
| 3 | 46368645 | 46368646 | rs41490645 |
| 3 | 46368544 | 46368545 | rs7637813 |
| 3 | 46948337 | 46948338 | rs7640397 |
| 4 | 122151853 | 122151854 | rs11938795 |
| 5 | 132490629 | 132490630 | rs10900809 |
| 5 | 96886184 | 96886185 | rs11135484 |
| 5 | 132490720 | 132490721 | rs11242115 |
| 5 | 96876036 | 96876037 | rs1230358 |
| 5 | 96886315 | 96886316 | rs13167902 |
| 5 | 96936802 | 96936803 | rs17684739 |
| 5 | 96936715 | 96936716 | rs2548224 |
| 5 | 132496821 | 132496822 | rs2548998 |
| 5 | 132491498 | 132491499 | rs2549005 |

| | | | |
|---|---|---|---|
| 5 | 132491372 | 132491373 | rs2549006 |
| 5 | 132491182 | 132491183 | rs2549007 |
| 5 | 132491072 | 132491073 | rs2549009 |
| 5 | 96935933 | 96935934 | rs2617434 |
| 5 | 132491187 | 132491188 | rs2706384 |
| 5 | 132496073 | 132496074 | rs2706385 |
| 5 | 132496994 | 132496995 | rs2706386 |
| 5 | 132490617 | 132490618 | rs960757 |
| 6 | 135497758 | 135497759 | rs13197384 |
| 7 | 28180608 | 28180609 | rs10255700 |
| 7 | 107168486 | 107168487 | rs112527862 |
| 7 | 22727025 | 22727026 | rs1800795 |
| 7 | 107168034 | 107168035 | rs257387 |
| 7 | 107168610 | 107168611 | rs4730220 |
| 7 | 107169203 | 107169204 | rs4730221 |
| 7 | 107169847 | 107169848 | rs4730222 |
| 7 | 107169661 | 107169662 | rs61014091 |
| 7 | 107169438 | 107169439 | rs62483623 |
| 7 | 107561129 | 107561130 | rs62483725 |
| 10 | 6050091 | 6050092 | rs10905668 |
| 10 | 6050129 | 6050130 | rs10905669 |
| 10 | 6070641 | 6070642 | rs10905713 |
| 10 | 6072046 | 6072047 | rs10905716 |
| 10 | 6072892 | 6072893 | rs10905718 |
| 10 | 86971004 | 86971005 | rs1240380 |
| 10 | 6072696 | 6072697 | rs41295061 |
| 10 | 6052733 | 6052734 | rs61839660 |
| 12 | 6383933 | 6383934 | rs28999107 |
| 12 | 111406413 | 111406414 | rs3742003 |
| 12 | 111599195 | 111599196 | rs695871 |
| 12 | 111412425 | 111412426 | rs7977752 |
| 13 | 42396586 | 42396587 | rs12874234 |
| 13 | 42396676 | 42396677 | rs12874725 |
| 13 | 42406741 | 42406742 | rs2324878 |
| 13 | 42393090 | 42393091 | rs34264781 |
| 13 | 42393136 | 42393137 | rs34700891 |
| 13 | 39655753 | 39655754 | rs3812882 |
| 13 | 39655819 | 39655820 | rs3812883 |
| 13 | 39655984 | 39655985 | rs3812885 |

| | | | |
|---|---|---|---|
| 13 | 39656078 | 39656079 | rs3812886 |
| 13 | 39656091 | 39656092 | rs3812887 |
| 13 | 39656156 | 39656157 | rs3812888 |
| 13 | 42405814 | 42405815 | rs7326472 |
| 13 | 39655586 | 39655587 | rs7327779 |
| 13 | 42392749 | 42392750 | rs7987414 |
| 13 | 42392875 | 42392876 | rs7987728 |
| 13 | 42397077 | 42397078 | rs7989519 |
| 13 | 39655430 | 39655431 | rs9548867 |
| 13 | 39655606 | 39655607 | rs9603589 |
| 14 | 68793572 | 68793573 | rs10443 |
| 14 | 68792944 | 68792945 | rs1051533 |
| 14 | 68780587 | 68780588 | rs11624323 |
| 14 | 68792688 | 68792689 | rs11847049 |
| 14 | 68792784 | 68792785 | rs11851414 |
| 14 | 68820765 | 68820766 | rs194742 |
| 14 | 68819434 | 68819435 | rs194743 |
| 14 | 68794754 | 68794755 | rs2236262 |
| 14 | 68780762 | 68780763 | rs56119720 |
| 14 | 68781230 | 68781231 | rs56232028 |
| 14 | 68793310 | 68793311 | rs57786342 |
| 14 | 68781539 | 68781540 | rs72731550 |
| 14 | 68793653 | 68793654 | rs72731554 |
| 14 | 68820939 | 68820940 | rs72731567 |
| 14 | 68780268 | 68780269 | rs76200426 |
| 19 | 10315745 | 10315746 | rs378395 |
| 22 | 30423639 | 30423640 | rs1061660 |
| 22 | 30423769 | 30423770 | rs1061664 |
| 22 | 21568166 | 21568167 | rs11089620 |
| 22 | 21630278 | 21630279 | rs11704601 |
| 22 | 30356952 | 30356953 | rs13053375 |
| 22 | 21568074 | 21568075 | rs140491 |
| 22 | 21568854 | 21568855 | rs140492 |
| 22 | 30356351 | 30356352 | rs17657653 |
| 22 | 30424088 | 30424089 | rs17658686 |
| 22 | 30426472 | 30426473 | rs2072158 |
| 22 | 21568614 | 21568615 | rs2266959 |
| 22 | 21630089 | 21630090 | rs3747093 |
| 22 | 30356706 | 30356707 | rs5994293 |

| | | | |
|---|---|---|---|
| 22 | 21629915 | 21629916 | rs710177 |
| 22 | 30396924 | 30396925 | rs740084 |
| 22 | 30397147 | 30397148 | rs757660 |
| 22 | 30397280 | 30397281 | rs887098 |
